# Beyond National Averages: Social Contact Patterns in the Upper Midwest

**DOI:** 10.64898/2026.11.13.26367651

**Authors:** Audrey Dorélien, Michala Carlson, Margo Wheatley, Lindsey Erickson, Keeley Morris, Kristin Sweet, Eva Enns

**Affiliations:** Department of Sociology University of Washington, 4100 Spokane Lane, Savery Hall 211, Mail code 353340, Seattle, Washington 98195, United States; Division of Health Policy and Management University of Minnesota, 420 Delaware St SE, MMC 729 Mayo 8729A Minneapolis, Minnesota 55455, United States; Minnesota Department of Health 625 N. Robert St., Saint Paul, MN, 55155 United States; Minnesota Department of Health 625 N. Robert St. Saint Paul, MN, 55155 United States; Division of Health Policy and Management University of Minnesota School of Public Health 420 Delaware St. SE, MMC 729, Minneapolis, MN, 55455, United States

**Keywords:** social contact patterns, contact matrices, mixing patterns, infectious disease transmission, regional variation, United States

## Abstract

**Background:** Social contact surveys measure how individuals interact with each other, which directly influences disease transmission dynamics. The COVID-19 pandemic substantially altered how people live and work, highlighting the need for up-to-date and sub-national estimates of social contact patterns to inform disease modeling and public health planning.

**Methods:** The Midwest Analytics & Disease Modeling Center (MADMC) conducted an opt-in social contact survey of upper Midwest residents, with about 60% of participants drawn from Minnesota. Participants reported all interpersonal contacts during the previous day, including the age of their contacts allowing for the analysis of mixing patterns. Detailed demographic information about the participants was also collected, allowing for additional subgroup analyses of contact patterns.

**Results:** Participants in our sample had an average of 6.8 contacts per day (IQR: 2.0 – 8.0), which is within the range of recent post-pandemic US national and European estimates. Contacts varied across demographic groups, with those attending work or school in-person reporting the highest number of contacts. Mixing across most age group pairs was a little lower than in comparable national survey except for younger age groups, who had substantially higher rates of contact with same-age peers. Differences in the contact matrices alone were associated with a 6.7% difference in estimated basic reproduction number(*R*_0_). Near the epidemic threshold, even this modest difference could meaningfully affect transmission dynamics. These effects may be amplified or attenuated by pathogen-specific age patterns of susceptibility and infectiousness, leading to larger or smaller differences in *R*_0_ estimates derived from national versus regional mixing patterns.

**Conclusions:** These findings provide empirical evidence on post-pandemic social contact patterns in the United States and highlight the potential implications of regional variation in social mixing for infectious disease transmission. The mean 6.8 daily contacts is consistent with evidence of persistent changes in daily routines following the pandemic. Incorporating contemporary, region-specific contact patterns into transmission models may improve their ability to capture geographic variation in transmission risk and inform epidemic forecasting.

## 1 Background

Understanding how people interact and come into contact with each other is vital to understanding infectious disease epidemiology. Erroneous assumptions about contact rates can lead to inaccuracies in mathematical model predictions, resulting in sub-optimal policies and interventions [1–3]. This was highlighted during the height of the COVID-19 pandemic, in which researchers and public health professionals had to advise on an effective response to the spread of a newly emergent, highly infectious, respiratory illness, but without localized information about population contact patterns or the extent to which policies like business closures and stay-at-home orders would modify those patterns [4].

Social contact patterns often vary by local context due to differences in sociodemographic features, such as age structure, racial and ethnic composition, household structure, work-force composition, as well as local policies and climate [2, 5, 6]. As a result, sub-national social contact surveys have proven to be essential to accurately measure local contact patterns, enabling more precise modeling of local infectious disease transmission dynamics and evaluation of control measures [6].

There is growing evidence that the COVID-19 pandemic has had long-lasting impacts on everyday life, including how people interact. The pandemic catalyzed sustained increases in work-from-home arrangements and hybrid work models have become the norm for many organizations [7]. Changes in where and how people work can have substantial impacts on social contact patterns, as workplace contacts historically have accounted for a large share of individuals’ total contacts [2, 8]. For instance, post pandemic contacts in the United Kingdom were 20% lower than pre-pandemic estimates [9]; and social contact changes in Europe were associated with a 20 to 50 % decline in *R*_0_ for a respiratory infectious disease compared to pre-pandemic period [8].

The Midwest Analytics and Disease Modeling Center’s Social Contact Survey (MADMC SCS) was developed to measure contemporary, post-pandemic contact patterns in the upper Midwest, addressing both the lack of sub-national contact data in the United States and the need for updated contact data to reflect post-pandemic behaviors. The survey collects information about contacts in household, work, school, and community settings, and combines detailed and aggregate reporting to better capture contact patterns of participants with both small and large numbers of contacts. We use these data to describe sociodemographic variation in daily number of contacts and to generate setting-specific, age-structured contact matrices. We also assess to what extent Upper Midwest contact patterns differ from national contact data and implication for infectious disease transmission.

## 2 Methods

### 2.1 Study population and sampling

The survey was administered online to an opt-in sample recruited by Ipsos, a well-established market research firm with extensive experience conducting opt-in health and behavioral surveys [10–13]. The study’s target population consisted of non-institutionalized individuals of all ages residing in Minnesota and neighboring states (North and South Dakota, Iowa, and Wisconsin). The survey was fielded online, in English, between September 14 and October 12, 2025. Survey respondents who provided a zip code located outside of these states were excluded from the study. Respondents were also excluded if they took longer than 48 hours to complete the survey or longer than 24 hours and crossing from weekday to weekend or vice versa. Participants were only allowed to complete the survey once.

Post-stratification weights were provided by the survey vendor to account for study design and to ensure sample representativeness of the upper Midwest region. Weights were generated to adjust survey responses to match upper Midwest region-specific population distribution by sex, age, race, state of residence, metropolitan status (as defined by the ACS/Office of Budget Management), education level, household income, and employment status, using an iterative proportional fitting (raking) procedure. Weights also adjusted for days of the week to weight each day of the week equally. Population targets were obtained from the 2023 American Community Survey [14]. To reduce the variance of the sample and the influence of outliers, weights were also trimmed and scaled to sum to the unweighted sample size of all qualified respondents.

### 2.2 Ethics statement

Participation in the MADMC SCS was voluntary. Consent was obtained from the participant or from the parent or legal guardian of participants under the age of 18 years. The consenting adult completed the survey on behalf of children 12 years and under, while children aged 13-17 years provided their assent and completed the contact modules of the survey independently after parent/legal guardian consent. The study was determined to be exempt from human subjects review by the University of Minnesota Institutional Review Board [ID: STUDY0024205].

### 2.3 Survey Design

The survey instrument can be found in Appendix A. We first asked participants about themselves and their household, including questions about age, sex, race/ethnicity, employment, level of education, household income, household size, marital status, and state of residence. Metropolitan status was determined from participant-reported residential zip code.

Participants were then asked about contacts they had on the previous day. A contact was defined as a person with whom the participant had an in-person conversation of at least five words or any physical contact (such as a handshake, hug, kiss, or contact sports). This definition was used to align with other social contact studies [15]. Contact information was obtained separately for four specific settings: household, work, school, and community. Participants were first asked about their household contacts. For each household contact, detailed information was collected, including but not limited to the contact’s age, sex, and race.

For work, school, and community contacts, participants were first asked to report the total number of contacts in each setting and then asked to list (using pseudonyms or nicknames) up to a maximum of 40 contacts in each setting. If a participant reported fewer than 10 contacts in a given setting, they were asked to provide detailed information for all of their listed contacts using questions similar to those for household members. For those reporting more than 10 contacts, detailed information was collected for a random sample of 10 of their listed contacts. Detailed information was limited to 10 contacts per setting to reduce survey fatigue. To ensure contact characteristics were still captured for individuals with large numbers of contacts, aggregate information was also collected by asking participants how their remaining contacts were distributed across age groups, allowing for potential imputation of other demographic characteristics. Community contacts also included a question about how the participant’s remaining contacts were distributed across different locations. Note that setting definitions were based on the participant’s relationship to that setting; a participant who worked in a school would report contacts with students and colleagues as work.

### 2.4 Statistical analyses

We first conducted descriptive analyses of the total number of contacts reported by participants (across all settings). Participant responses were top-coded (an upward bound to the total number of contacts) at 40 total contacts, similar to other social contact surveys [2, 16, 17], to reduce the influence of outliers on analysis results. We examined the distribution of the total number of contacts to assess whether there was evidence of heaping at multiples of 5 or 10, which could indicate participants were loosely estimating rather than re-calling exact number of contacts. We evaluated how the number of reported contacts varied by participant characteristics, such as sex, age, household size, school attendance (in-person, online, no school), work type (in-person vs. not in-person), household income, residence in a metro area, and day of the week contacts were reported for (weekend vs. weekday). We used negative binomial regression models to estimate contact rate ratios (RRs) associated with these factors, in both univariate (unadjusted) and multivariate (adjusted) analyses to identify statistically significant differences [13, 16]. All analyses were conducted using sample weights. In sensitivity analysis, we ran regression models without top-coding contacts, and we also evaluated the performance of Zero Inflated Poisson and Zero Inflated Negative Binomial regression models in fitting contact data.

#### 2.4.1 Mixing patterns

Mixing matrices are used to describe how different groups interact with one another and the extent to which contacts are concentrated within groups (homophily or assortativity) versus across groups. Because age is closely linked with key social contexts such as school, work, and household composition [1], we constructed age-structured mixing matrices to reflect how participants of different ages interact with contacts of different ages. We constructed an overall age-structured mixing matrix, as well as separate, setting-specific age-structured matrices for household, work, school, and community. In the contact data, the maximum number of contacts a participant could have was capped at 40 by location.

We constructed age-structured contact matrices by first assigning participants and their contacts to 10-year age groups starting at age 0, with ages 80 years and older being the oldest age group. Each cell of the age-structured contact matrix, *M_ij_*, represents the *weighted* mean number of daily contacts between participants in age group *i* and their contacts in age group *j*. Each participant *t* is associated with a sample weight, *w_t_*, that adjusts their characteristics, including age, sex, race, and day of the week, to construct a population-representative estimate. Specifically, we calculate *M_ij_* as:

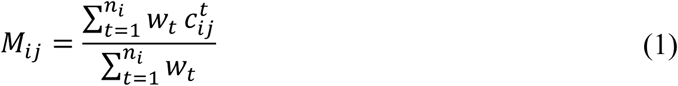

where *c^t^_ij_* is the number of contacts reported by participant t in age group i with contacts in age group *j*; *w_t_* is the sample weight for participant *t*; and *n_i_* is the unweighted number of participants in age group *i*. For setting-specific age-structured matrices, the number of contacts *c^t^_ij_* is restricted to those reported in the setting of interest.

For each age-structured contact matrix, we calculated the Q-index, a measure of homophily or assortativeness [13, 16, 18]. A value of *Q* = 0 indicates homogeneous mixing, whereas *Q >* 0 indicates assortative mixing, with *Q* = 1 corresponding to complete assortative mixing, meaning that people only mix with others within the same age group. Finally, values of *Q <* 0 indicate disassortative mixing, meaning that participants are more likely to mix with people outside of their age group compared to people within it. We generate the Q-index by first normalizing *M_ij_* (turning counts into proportions):

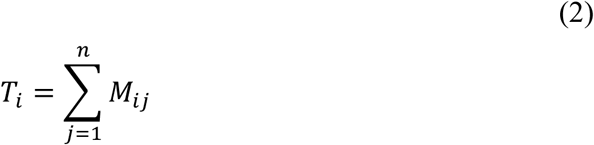

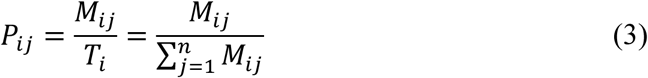

where *P_ij_* is the proportion of contacts of participants in age group *i* with contacts in age group *j*, and *n* is the number of age groups. The Q-index is then calculated as:

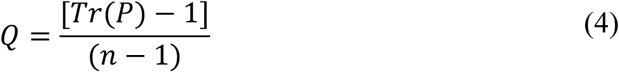

where *Tr*(*P* ) is the trace of the proportional mixing matrix *P*.

Because of differences in reporting, the matrices that we generated are not symmetric/reciprocal [16, 19]. Symmetric versions of our contact matrices are provided in Appendix C.

#### 2.4.2 Comparing Mixing Patterns and Implications for Disease Transmission

We compared the MADMC SCS age-structured contact matrices with the recently collected and nationally representative Epistorm-Mix study [13]. Before comparison, all matrices were harmonized to use the same age-group definitions. The Epistorm-Mix study included multiple survey waves during the spring, fall, and winter. To reduce differences that might arise due to seasonality, MADMC SCS results were only compared to the Epistorm-Mix fall 2024 survey results.

We summarized cell-level agreement between MADMC SCS and Epistorm-Mix contact matrices using Pearson’s correlation coefficient and Spearman’s rank correlation coefficient. To assess the magnitude and direction of the differences, we also calculated the cell-level difference between the MADMC contact matrix and the Epistorm-Mix matrix. We visualized these differences as heatmaps to identify whether the greatest differences occurred along the main diagonal, between adjacent age groups, or in substantively important off-diagonal cells, such as adult-child contacts. Finally, to compare transmission potential, we estimated the basic reproduction number (*R*_0_) using both the Epistorm-Mix and MADMC contact matrices for a respiratory pathogen assuming uniform susceptibility across age groups [20]. For each matrix, we constructed a next-generation matrix (NGM) and calculated its dominant eigenvalue, which is proportional to *R*_0_ when transmission parameters are held constant. The resulting proportional *R*_0_ were then compared to assess differences in transmission potential attributable solely to differences in the contact matrices.

All analyses were conducted in R version 4.6.0. We will make the code and data available on GitHub.

## 3 Results

### 3.1 Survey sample characteristics

The MADMC SCS analytic sample consisted of 1,009 individuals reporting 5,806 unique contacts (when the maximum number of contacts was top-coded at 40). Sixty percent (n = 612) of the participants resided in Minnesota and the remaining 40% resided in surrounding states. Slightly more than half (53%) of our sample was female, and the majority were from metro areas (78%). Our sample was primarily White (90%); therefore, we did not stratify results by race and ethnicity. All days of the week were represented in the survey; before applying weights, approximately 23% of participants reported contacts that occurred on a Saturday or Sunday.

### 3.2 Number of contacts

The distribution of reported total contacts (across all settings) was right-skewed (Figure 1) and did not show evidence of heaping at multiples of 5 and 10. In the sample, the mean number of contacts was 6.84 and the median was 4.00 (IQR: 2.0-8.0). Without top-coding total contacts at 40, the mean shifted to 8.68 while the median remained at 4.00 (IQR: 2.0-8.0).

**Figure 1.**
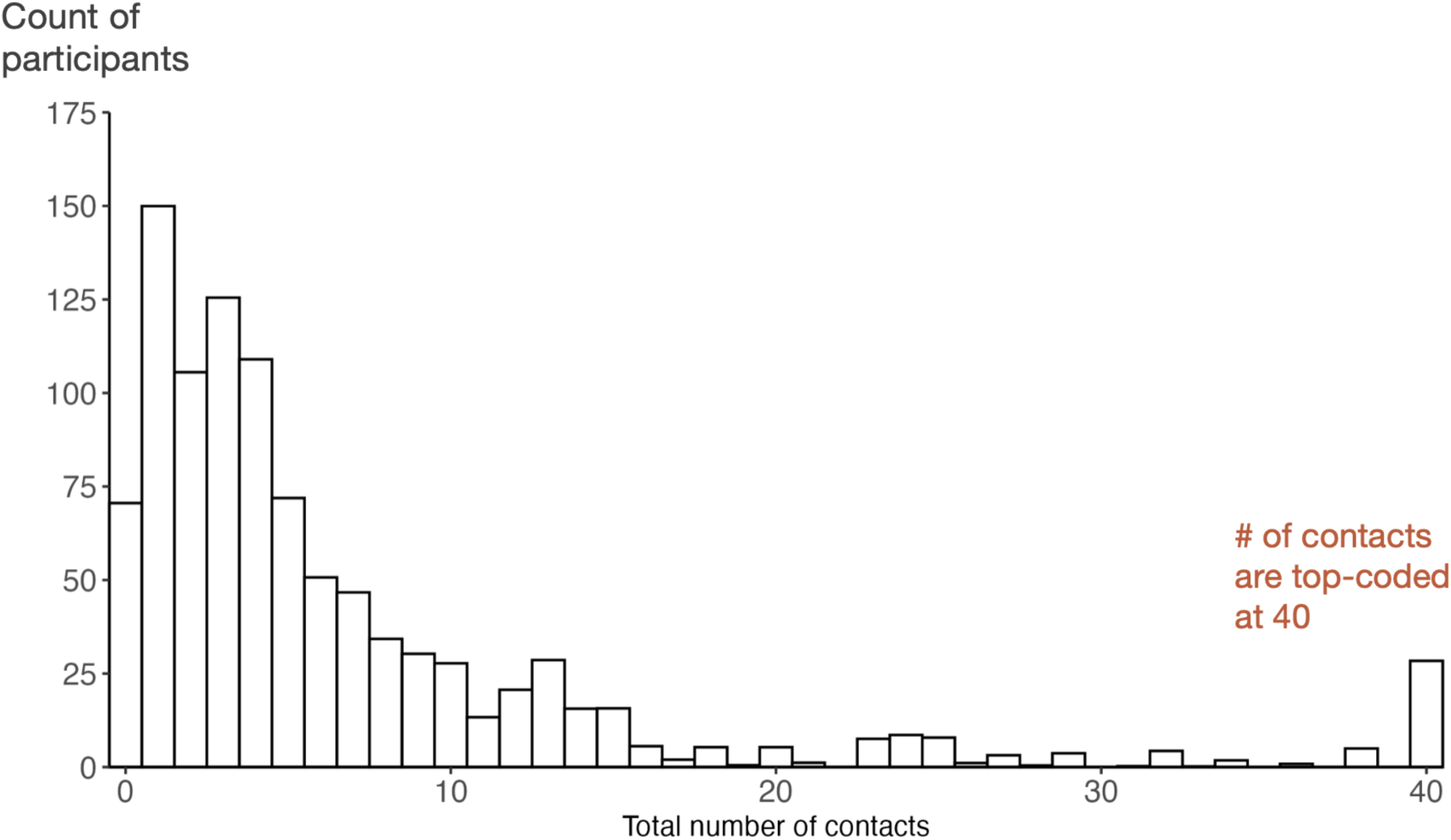
Histogram showing the distribution of the total number of daily contacts. Mean is 6.8 and median is 4.0. Maximum number of contacts is top-coded at 40.

### 3.3 Correlates of contacts

Table 1 summarizes the number of reported contacts across sociodemographic groups, including the mean, median, and interquartile range. Table 2 presents unadjusted and adjusted rate ratios from negative binomial regression models with reference categories as indicated in the table.

**Table 1.** Number of contacts by participant characteristics. Summary statistics (mean, median, and interquartile range [IQR]) for the number of contacts are reported for each sociodemographic group.

|  |  | Descriptive Results |  |  |  |
|---|---|---|---|---|---|
| Variable | Level | N (%) | Mean | Median | IQR |
| Overall |  | 1009 (100.0) | 6.8 | 4.0 | 2.0 – 8.0 |
| **Sex** | **Female** | **533 (52.8)** | **6.0** | **3.0** | **1.0 – 7.0** |
|  | Male | 476 (47.2) | 7.7 | 5.0 | 2.0 – 9.0 |
| Age group | 0–4 | 61 (6.0) | 6.5 | 5.0 | 3.0 – 6.0 |
|  | 5–9 | 58 (5.7) | 11.3 | 5.0 | 3.0 – 15.0 |
|  | 10–14 | 58 (5.7) | 9.7 | 7.0 | 3.0 – 14.0 |
|  | 15–19 | 38 (3.8) | 16.3 | 8.0 | 5.0 – 38.0 |
|  | 20–24 | 24 (2.4) | 7.8 | 5.0 | 2.0 – 12.0 |
|  | **25–29** | **48 (4.8)** | **9.9** | **6.0** | **4.0 – 10.0** |
|  | 30–34 | 47 (4.7) | 8.1 | 5.0 | 3.0 – 11.0 |
|  | 35–39 | 85 (8.4) | 6.3 | 5.0 | 2.0 – 8.0 |
|  | 40–44 | 78 (7.7) | 5.0 | 4.0 | 2.0 – 7.0 |
|  | 45–49 | 66 (6.5) | 5.9 | 4.0 | 2.0 – 9.0 |
|  | 50–54 | 67 (6.6) | 5.1 | 4.0 | 1.0 – 5.0 |
|  | 55–59 | 59 (5.8) | 4.3 | 2.0 | 1.0 – 5.0 |
|  | 60–64 | 88 (8.7) | 4.9 | 2.0 | 1.0 – 5.0 |
|  | 65–69 | 81 (8.0) | 3.4 | 2.0 | 1.0 – 5.0 |
|  | 70–74 | 64 (6.3) | 2.6 | 1.0 | 1.0 – 3.0 |
|  | 75+ | 87 (8.6) | 3.0 | 2.0 | 1.0 – 5.0 |
| **Household size** | **1** | **176 (17.4)** | **4.4** | **3.0** | **0.0 – 5.0** |
|  | 2 | 305 (30.2) | 5.3 | 2.0 | 1.0 – 6.0 |
|  | 3 | 212 (21.0) | 5.8 | 3.0 | 2.0 – 6.0 |
|  | 4 | 189 (18.7) | 8.6 | 6.0 | 3.0 – 12.0 |
|  | 5 | 80 (7.9) | 8.3 | 5.0 | 4.0 – 10.0 |
|  | 6 or more | 47 (4.7) | 13.8 | 9.0 | 6.0 – 20.0 |
| **School** | **No school** | **817 (81.0)** | **5.2** | **3.0** | **1.0 – 7.0** |
|  | Online/remote | 44 (4.4) | 10.1 | 5.0 | 3.0 – 12.0 |
|  | In person school | 148 (14.7) | 13.1 | 8.0 | 4.0 – 17.0 |
| **Work** | **In person work** | **232 (23.0)** | **9.5** | **8.0** | **4.0 – 12.0** |
|  | No in person work | 777 (77.0) | 5.7 | 3.0 | 1.0 – 6.0 |
| **Household Income** | **Less than \$10,000** | **78 (7.7)** | **5.9** | **3.0** | **0.0 – 5.0** |
|  | \$10,000 to \$24,999 | 105 (10.4) | 4.4 | 2.0 | 1.0 – 6.0 |
|  | \$25,000 to \$49,999 | 234 (23.2) | 6.9 | 4.0 | 1.0 – 8.0 |
|  | \$50,000 to \$74,999 | 203 (20.1) | 4.2 | 3.0 | 1.0 – 5.0 |
|  | \$75,000 to \$99,999 | 152 (15.1) | 6.0 | 4.0 | 2.0 – 7.0 |
|  | \$100,000 to \$149,999 | 139 (13.8) | 7.8 | 5.0 | 2.0 – 9.0 |
|  | \$150,000 or more | 98 (9.7) | 9.3 | 6.0 | 3.0 – 12.0 |
| **Metro Status** | **Metro** | **782 (77.5)** | **7.1** | **4.0** | **2.0 – 8.0** |
|  | Non-Metro | 227 (22.5) | 6.1 | 4.0 | 2.0 – 9.0 |
| **Type of Day** | **Weekend** | **232 (23.0)** | **6.8** | **3.0** | **2.0 – 6.0** |
|  | Weekday | 777 (77.0) | 6.9 | 4.0 | 2.0 – 9.0 |

*N and % represent unweighted sample counts and category percentages; means, medians, and interquartile ranges [IQR] are weighted. IQR reported as 25th-75th percentiles (Q1-Q3).*

**Table 2.**
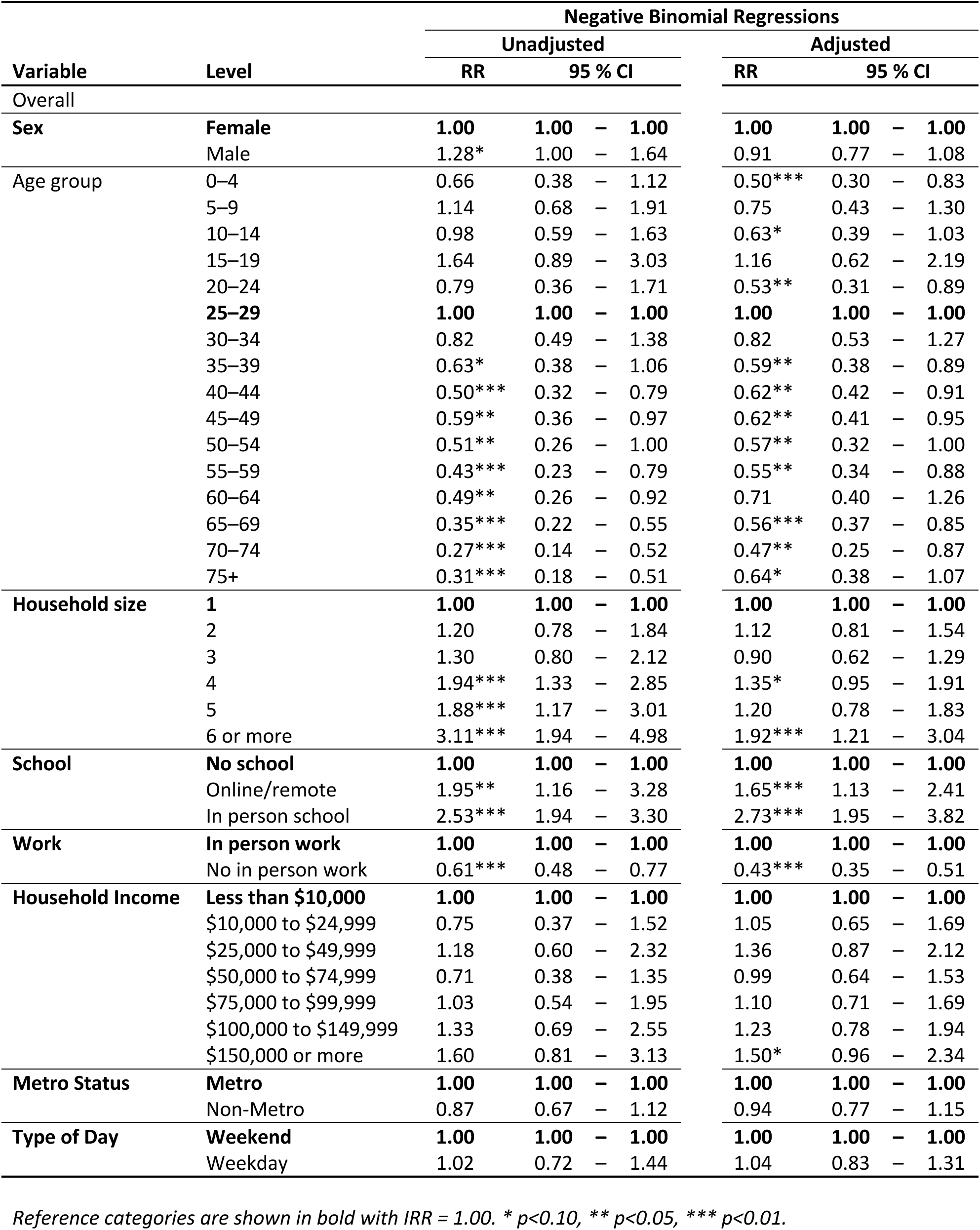
Unadjusted and adjusted rate ratios (RR) for contact counts by sociodemographic factors.

|  |  | Negative Binomial Regressions |  |  |  |  |  |  |  |
|---|---|---|---|---|---|---|---|---|---|
|  |  | Unadjusted |  |  |  | Adjusted |  |  |  |
| Variable | Level | RR | 95 % CI |  |  | RR | 95 % CI |  |  |
| Overall |  |  |  |  |  |  |  |  |  |
| **Sex** | **Female** | **1.00** | **1.00** | **–** | **1.00** | **1.00** | **1.00** | **–** | **1.00** |
|  | Male | 1.28* | 1.00 | – | 1.64 | 0.91 | 0.77 | – | 1.08 |
| Age group | 0–4 | 0.66 | 0.38 | – | 1.12 | 0.50*** | 0.30 | – | 0.83 |
|  | 5–9 | 1.14 | 0.68 | – | 1.91 | 0.75 | 0.43 | – | 1.30 |
|  | 10–14 | 0.98 | 0.59 | – | 1.63 | 0.63* | 0.39 | – | 1.03 |
|  | 15–19 | 1.64 | 0.89 | – | 3.03 | 1.16 | 0.62 | – | 2.19 |
|  | 20–24 | 0.79 | 0.36 | – | 1.71 | 0.53** | 0.31 | – | 0.89 |
|  | **25–29** | **1.00** | **1.00** | **–** | **1.00** | **1.00** | **1.00** | **–** | **1.00** |
|  | 30–34 | 0.82 | 0.49 | – | 1.38 | 0.82 | 0.53 | – | 1.27 |
|  | 35–39 | 0.63* | 0.38 | – | 1.06 | 0.59** | 0.38 | – | 0.89 |
|  | 40–44 | 0.50*** | 0.32 | – | 0.79 | 0.62** | 0.42 | – | 0.91 |
|  | 45–49 | 0.59** | 0.36 | – | 0.97 | 0.62** | 0.41 | – | 0.95 |
|  | 50–54 | 0.51** | 0.26 | – | 1.00 | 0.57** | 0.32 | – | 1.00 |
|  | 55–59 | 0.43*** | 0.23 | – | 0.79 | 0.55** | 0.34 | – | 0.88 |
|  | 60–64 | 0.49** | 0.26 | – | 0.92 | 0.71 | 0.40 | – | 1.26 |
|  | 65–69 | 0.35*** | 0.22 | – | 0.55 | 0.56*** | 0.37 | – | 0.85 |
|  | 70–74 | 0.27*** | 0.14 | – | 0.52 | 0.47** | 0.25 | – | 0.87 |
|  | 75+ | 0.31*** | 0.18 | – | 0.51 | 0.64* | 0.38 | – | 1.07 |
| **Household size** | **1** | **1.00** | **1.00** | **–** | **1.00** | **1.00** | **1.00** | **–** | **1.00** |
|  | 2 | 1.20 | 0.78 | – | 1.84 | 1.12 | 0.81 | – | 1.54 |
|  | 3 | 1.30 | 0.80 | – | 2.12 | 0.90 | 0.62 | – | 1.29 |
|  | 4 | 1.94*** | 1.33 | – | 2.85 | 1.35* | 0.95 | – | 1.91 |
|  | 5 | 1.88*** | 1.17 | – | 3.01 | 1.20 | 0.78 | – | 1.83 |
|  | 6 or more | 3.11*** | 1.94 | – | 4.98 | 1.92*** | 1.21 | – | 3.04 |
| **School** | **No school** | **1.00** | **1.00** | **–** | **1.00** | **1.00** | **1.00** | **–** | **1.00** |
|  | Online/remote | 1.95** | 1.16 | – | 3.28 | 1.65*** | 1.13 | – | 2.41 |
|  | In person school | 2.53*** | 1.94 | – | 3.30 | 2.73*** | 1.95 | – | 3.82 |
| **Work** | **In person work** | **1.00** | **1.00** | **–** | **1.00** | **1.00** | **1.00** | **–** | **1.00** |
|  | No in person work | 0.61*** | 0.48 | – | 0.77 | 0.43*** | 0.35 | – | 0.51 |
| **Household Income** | **Less than \$10,000** | **1.00** | **1.00** | **–** | **1.00** | **1.00** | **1.00** | **–** | **1.00** |
|  | \$10,000 to \$24,999 | 0.75 | 0.37 | – | 1.52 | 1.05 | 0.65 | – | 1.69 |
|  | \$25,000 to \$49,999 | 1.18 | 0.60 | – | 2.32 | 1.36 | 0.87 | – | 2.12 |
|  | \$50,000 to \$74,999 | 0.71 | 0.38 | – | 1.35 | 0.99 | 0.64 | – | 1.53 |
|  | \$75,000 to \$99,999 | 1.03 | 0.54 | – | 1.95 | 1.10 | 0.71 | – | 1.69 |
|  | \$100,000 to \$149,999 | 1.33 | 0.69 | – | 2.55 | 1.23 | 0.78 | – | 1.94 |
|  | \$150,000 or more | 1.60 | 0.81 | – | 3.13 | 1.50* | 0.96 | – | 2.34 |
| **Metro Status** | **Metro** | **1.00** | **1.00** | **–** | **1.00** | **1.00** | **1.00** | **–** | **1.00** |
|  | Non-Metro | 0.87 | 0.67 | – | 1.12 | 0.94 | 0.77 | – | 1.15 |
| **Type of Day** | **Weekend** | **1.00** | **1.00** | **–** | **1.00** | **1.00** | **1.00** | **–** | **1.00** |
|  | Weekday | 1.02 | 0.72 | – | 1.44 | 1.04 | 0.83 | – | 1.31 |

*Reference categories are shown in bold with IRR = 1.00. \* p<0.10, \*\* p<0.05, \*\*\* p<0.01.*

Men reported more contacts than women (RR = 1.28) in the unadjusted results, with a mean of 7.7 (IQR: 2.0–9.0) contacts per day compared to 6.0 (IQR: 1.0–7.0) among women. This difference was marginally statistically significant in the unadjusted model. The men in the sample were also more likely to work in person than the women, suggesting that work-related exposure may partly explain the observed difference. After adjusting for covariates, men had a lower estimated contact rate than women (RR = 0.91), but this difference was not statistically significant.

The average number of contacts reported per day varied substantially by age. Contacts were highest among adolescents and young adults, peaking at a mean of 16.3 (IQR: 5.0 – 38.0) daily contacts among participants aged 15 – 19 years. Children aged 5 – 9 and 10 – 14 years also reported relatively high contact rates, with means of 11.3 (IQR: 3.0 – 15.0) and 9.7 (IQR: 3.0 – 14.0) contacts per day, respectively. Contact rates declined for participants from older age groups, falling from an average of 8–10 contacts per day among participants in their 20s and early 30s to about an average of 5–6 contacts per day among participants aged 35 - 55. Adults aged 55 and older reported the fewest contacts, with means below 5 (IQR: 1.0-5.0) contacts per day. Participants aged 70 - 75 years had the lowest mean number of daily contacts (2.6; IQR: 1.0–3.0).

For participants aged 5 – 19, most reported contacts took place at school. After adjustments for covariates, including school attendance and in-person work, age group differences were attenuated. Participants aged 15-19 remained the most connected (RR = 1.16), followed by those aged 25-30 (Table 2). In contrast, most other age groups converged to a similar mean number of contacts per day, and the pronounced decline in contact rates at older ages was no longer observed.

Reported contacts increased with household size. Participants living alone averaged 4.4 contacts per day, while those in two- or three-person households averaged 5.3 to 5.8 contacts per day. Contact rates were higher in larger households, rising to 8.6 contacts per day among participants in four-person households and 13.8 among those in households with six or more members. Relative to participants living alone, the difference was statistically significant only for households with more than four members. After controlling for covariates, only those in households with six or more people had statistically significantly more contacts than those living alone (RR = 1.92).

Participants in households with annual incomes above $100,000 reported more contacts (mean = 7.8-9.3) than those in lower-income households, particularly those earning less than $25,000 (mean = 5.1; IQR: 0.0-5.0) or between $50,000 and $75,000 (mean = 4.2; IQR: 1.0-5.0). Contact rates did not increase monotonically with household income. Differences by income were not statistically significant in both unadjusted or adjusted models (p > 0.05). Students had higher contact rates than non-students. Students attending school in-person reported an average 13.1 contacts per day (IQR:4.0-17.0) compared to online students 10.1 contacts per day (IQR:3.0-12.0) and non-students who reported 5.2 contacts per day (IQR:1.0-7.0). In our sample, online-students were older and were more likely to work in-person.

Work modality also impacted reported daily contacts. Those who work in-person at least some of the time reported an average of 9.5 contacts per day (IQR: 4.0-12.0) compared to 5.7 (IQR: 1.0-6.0) contacts per day for those who do not work or who work remotely. This difference remains after controlling for covariates.

Contact rates were somewhat lower among participants in non-metro areas than among those in metro areas. Non-metro participants reported an average of 6.2 contacts per day (IQR: 2.0–9.0), compared with 7.1 contacts per day among metro participants (IQR: 2.0–8.8). This difference was not statistically significant in the unadjusted model (RR = 0.87), and controlling for covariates did not materially change the results (RR = 0.94). Because metro participants were more likely to report more than 40 contacts, the metro–non-metro difference appeared larger when contact counts were analyzed without top-coding (see Supple-mental Materials Appendix B).

Reported contacts were similar on weekdays and weekends. Participants reported an average of 6.9 contacts per day on weekdays (IQR: 2.0-9.0) and 6.8 contacts per day on weekends (IQR: 2.0-6.0). When contact counts were analyzed without top-coding, weekend contacts appeared much higher than weekday contacts, however, this difference was no longer statistically significant after adjusting for covariates.

### 3.4 Age-Structured Mixing Patterns

Figure 2 presents the overall age-structured contact matrix (10-year age groups), along with setting-specific contact matrices and the corresponding age-assortativity (Q-index) for each. We also calculated the share of age-specific contacts in each cell reported to be with household members (Figure 2F).

**Figure 2.**
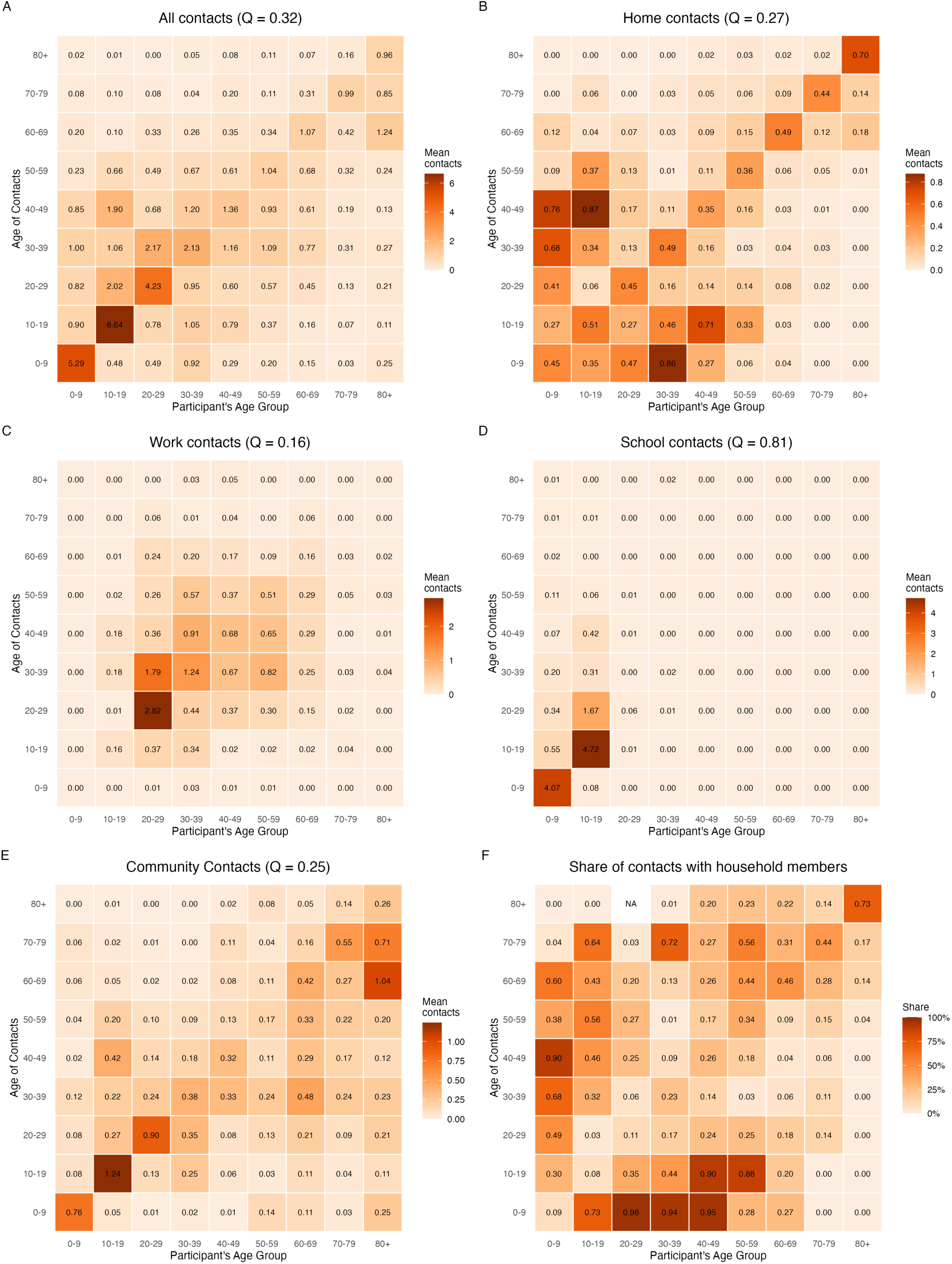
Age-Structured Contact Matrices by Setting. Q-index calculation for work excludes children and elderly. Q-index calculation for school excludes those older than 19 years and younger than 5 years.

Across most age groups, the greatest amount of mixing took place between participants and similarly aged contacts. The exception was middle aged participants (40-49) being slightly more likely to interact with children aged 10-19. Children and young adults exhibited the highest levels of age-assortative mixing, averaging 4 to 7 same age contacts per day. Among participants younger than 19, these contacts occurred primarily in school (Figure 2D) and in community settings (Figure 2E). In contrast, among young adults, most assortative contacts took place at work (Figure 2C). Participants aged 50 and above had about one contact per day within their age group.

In the home setting, children and adolescents primarily mixed with siblings, while adults mixed with their similarly aged partners or other household members. However, the highest number of home contacts reflected inter-generational mixing, particularly between parents aged 30-49 and children aged 0-19. The lower diagonal represents parents interacting with children, while the upper diagonal represents children interacting with their parents (Figure 2B).

Most workplace contacts occurred among adults aged 20-69, which is consistent with the working age population. The levels of workplace mixing occurred within the 20-29 and 30-39 age groups and between these two adjacent age groups (Figure 2C). The overall workplace Q-index is low (0.09) suggesting homogeneous mixing; we also see evidence of homogeneous mixing when we exclude children and elderly from the analysis (Q-index = 0.14). In contrast, although the overall Q-index for school is also very low (Q-index = 0.09), restricting the analysis to children under age of 19 results in a very high level of homophily (Q-index = 0.83) (Figure 2D). At school, children averaged 4-5 contacts per day with other children.

Community contacts also exhibited a strong age assortative profile (Figure 2E). It is in community settings that the elderly mixed with other age groups the most, especially with adjacent age groups. Children under 10 had very few community contacts with other age groups.

### 3.5 Comparison with Epistorm-Mix Matrices

The Epistorm-Mix data we use had 980 participants with 7,492 contacts. The maximum number of contacts contributed by a single participant was similar across the two datasets (MADMC SCS: 77; Epistorm-Mix: 67), suggesting that extreme values were unlikely to substantially influence the age-structured contact matrices. In Epistorm-Mix, 28 participants reported 40 or more contacts, including 9 who reported more than 49 con-tacts. In comparison, MADMC SCS included 16 participants with 40 or more contacts, of whom only 3 reported more than 49 contacts. We found a strong statistical correspondence between the national Epistorm-Mix matrix and the MADMC SCS contact matrix (Pearson *r*= 0.92; Spearman *ρ*= 0.90; R^2^= 0.84). The rank relationship was even stronger when focusing exclusively on the diagonal elements which capture age-assortative mixing (Pearson *r*= 0.92; Spearman *ρ*= 0.97). Despite the overall similarity, the Epistorm-Mix matrix generally exhibited higher contact rates than the MADMC SCS matrix across most age-group pairings. The primary exception was among younger age groups (*<*30 years), where MADMC SCS participants reported substantially higher rates of contact within their own age group, reflected in the stronger age-assortative mixing along the matrix diagonal (Figure 3).

**Figure 3.**
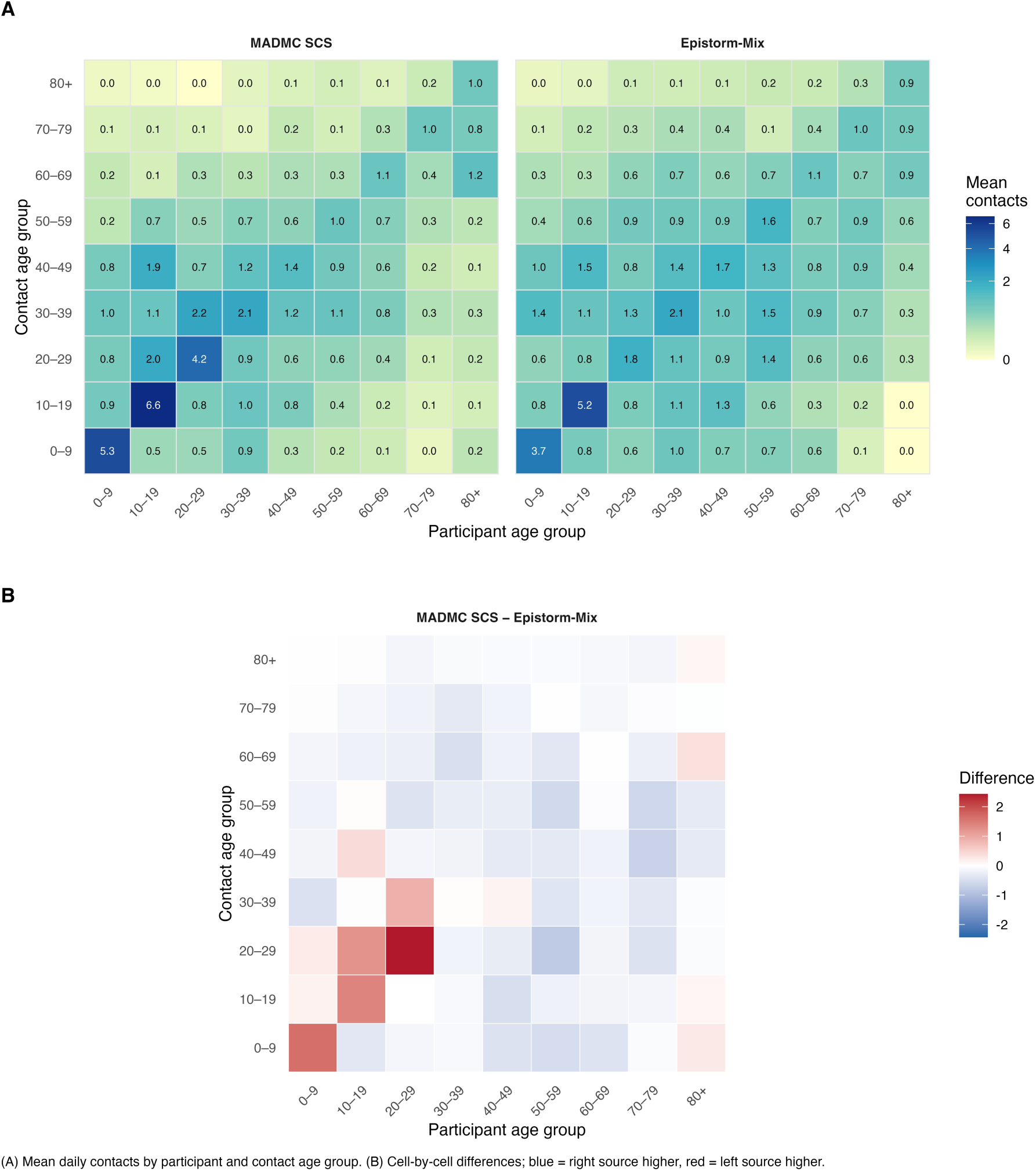
Comparing age-structured contact matrices for the fall waves of the MADMC SCS and Epistorm-Mix.

The leading eigenvalue of the MADMC SCS contact matrix was 8.84, compared with 8.28 for the Epistorm matrix. The resulting relative *R*_0_ (MADMC/Epistorm) was 1.067, indicating that the MADMC SCS contact matrix yielded an *R*_0_ 6.7% higher than that obtained using the Epistorm contact matrix, holding all other transmission parameters constant.

## 4 Discussion and Conclusion

In the fall of 2025, the MADMC Social Contact Survey recorded an average of 6.8 contacts per day (IQR: 2.0–8.0) among participants in Minnesota and surrounding upper Midwest states. When we remove the upward bound on number of contacts, the reported mean was 8.7 contacts per day (IQR: 2.0-8.0). Both estimates fall within the range of recent post-pandemic European estimates of mean daily contacts, including 6.0 contacts per day in Switzerland, 6.5 contacts per day in the United Kingdom, 6.7 contacts per day in Belgium, and 9.9 in the Netherlands [8]. Our top-coded MADMC SCS estimates were lower than the national US-focused Epistorm-Mix study collected in fall 2024, which estimated 7.4 contacts per day (IQR: 2.0–9.0) nationally and 8.0 contacts per day (IQR: 3.0–10.0) in their Midwestern regional subsample [13]. However, after removing the top-coding our estimate were slightly higher.

Despite lower total daily contacts than contemporary US national estimates, the Midwest sub-national mixing patterns by age exhibited greater within-age-group transmission potential among children, adolescents, and young adults. These highly connected age groups can form a transmission “core” that can disproportionately influence epidemic dynamics. Correspondingly, this difference in age-based mixing resulted in an estimated 6.7% higher *R*_0_ than that implied by national contact patterns. Although modest in relative terms, a difference of this magnitude may still be epidemiologically important when transmission is near the epidemic threshold, where relatively small changes in *R*_0_ can substantially alter epi-demic growth rates. Furthermore, the 6.7% relative difference in *R*_0_ is not fixed and could increase or decrease based on interaction between mixing patterns and specific infectious pathogens age pattern of susceptibility and infectiousness. The observed differences may reflect underlying geographic variation as well as differences in the study period and sampling methodology, particularly the contrast between the probability-based Epistorm-Mix sample and the MADMC SCS opt-in sample. There were also differences in how MADMC SCS and Epistorm-Mix treated participants with large number of contacts. As additional national and sub-national data become available, it will be important to assess whether these patterns persist.

Although comparable pre-pandemic estimates of social contact patterns are not avail-able for the Upper Midwest or the United States nationally, MADMC contact rates were approximately half of those reported in historical European studies [2]. This difference is consistent with broader evidence that the pandemic produced persistent changes in daily routines, including increased work-from-home arrangements.

School and work settings were the primary drivers of social contact patterns. Consistent with prior research, participants attending school or working in person reported substantially more daily contacts than those who did not, and age groups most engaged in these activities (15-19 and 24-29 years) had the highest contact rates [13]. Although individuals in online school also reported high contact rates, most were employed adults whose contacts occurred predominantly in workplaces (Appendix D1). These findings suggest that continued changes in work arrangements, particularly shifts toward or away from in-person work, could meaningfully affect population-level contact rates and transmission potential. Higher household income was also associated with greater contact rates, a pattern largely explained by more school-related contacts, including daycare, camp, and after-school care, further underscoring the central role of schools in shaping social mixing (Appendix D2). At the same time, substantial intergenerational mixing in community settings, particularly between older adults and younger age groups, suggests that community-based interventions may be important for reducing exposure among susceptible older adults.

Additional research is needed to investigate the potential impact of sampling approaches and survey instrument design. The MADMC social contact survey used an opt-in sample, providing a source of potential bias in the study population. The post-stratification weights ensured that our sample aligned with key demographic characteristics of the population, but some unobserved differences may still remain. We were reassured that our findings were consistent with known mixing patterns and that our results were largely consistent with data from Epistorm-Mix, the nationally representative survey that utilized probability sampling.

A second limitation is the reliance on self-reported contact data, which can be subject to reporting bias. Standard contact survey instruments can be lengthy and burdensome to complete, particularly for individuals with many contacts, and survey fatigue may lead some respondents to under report their true number of interactions. To assess one potential source of reporting bias, we examined the distribution of reported contacts across all contact modules (home, school, work, and other) for evidence of “heaping” at multiples of 5 and 10. The resulting histograms showed little evidence of systematic rounding, suggesting that this source of reporting bias was limited (Appendix E). Nevertheless, further work is needed to identify methods that improve contact ascertainment and reduce under reporting, particularly among highly connected individuals. Such advances would support more accurate estimates of contact patterns and strengthen inferences about their implications for disease transmission.

In conclusion, contact patterns from the Upper Midwest were similar to national estimates. However, there were key differences such as higher rates of assortative mixing among participants younger than 30 years of age in the Upper Midwest. These data are important for better understanding post-pandemic social contact behaviors and potential regional differences that exist.

## Data Availability

All data produced in the present work are available online in a GitHub repository: https://github.com/MADM-C/scs-fall-manuscript

## Ethics Approval declaration

This study was performed in compliance with relevant laws, regulatory frameworks and guidelines where the research took place.

The **University of Minnesota Institutional Review Board** granted an exemption. The study was determined to be exempt from human subjects review by the **University of Minnesota Institutional Review Board [ID: STUDY0024205].** The study met the following criteria: Research that only includes interactions involving educational tests (cognitive, diagnostic, aptitude, achievement), survey procedures, interview procedures, or observation of public behavior (including visual or auditory recording) if at least one of the following criteria is met: (ii) Any disclosure of the human subjects’ responses outside the research would not reasonably place the subjects at risk of criminal or civil liability or be damaging to the subjects’ financial standing, employability, educational advancement, or reputation.

## Consent for Publications

Not applicable

## Availability of Data and Materials

The participant and contact datasets used during the current study are available from the corresponding author on reasonable request. The data will be made available in a GitHub Repository.

## Author contributions

A.D., E.E., and K.S. conceptualized the study. A.D., E.E., M.C., M.W., L.E, K.M. contributed to the design of the survey instrument and analysis plan. A.D., M.C., and M.W. supported data collection. A.D. oversaw the data analysis, which was conducted by M.C. A.D. led the writing of the manuscript with input from all authors. All authors provided critical feedback and helped shape the research, analysis and manuscript.

## Acknowledgments

We thank IPSOS, specifically Sergei Rodkin and Ying Wang for their support with survey design and implementation. We would like to thank Dr. Ayesha Mahmud and Dr. Dennis Feehan for their feedback on the survey design and Mingze Li for code review.

## References

[1] Jacco Wallinga, Peter Teunis, and Mirjam Kretzschmar. Using Data on Social Contacts to Estimate Age-specific Transmission Parameters for Respiratory-spread Infectious Agents. American Journal of Epidemiology, 164(10):936–944, November 2006. ISSN 0002-9262. doi: 10.1093/aje/kwj317. URL https://academic.oup.com/aje/article/164/10/936/162511. tex.ids: wallinga using 2006-1.

[2] Joël Mossong, Niel Hens, Mark Jit, Philippe Beutels, Kari Auranen, Rafael Mikolajczyk, Marco Massari, Stefania Salmaso, Gianpaolo Scalia Tomba, Jacco Wallinga, Janneke Heijne, Malgorzata Sadkowska-Todys, Magdalena Rosinska, and W. John Edmunds. Social Contacts and Mixing Patterns Relevant to the Spread of Infectious Diseases. PLoS Medicine, 5(3):e74, March 2008. ISSN 1549-1676. doi: 10.1371/journal.pmed.0050074. URL https://dx.plos.org/10.1371/journal.pmed.0050074.

[3] Kiesha Prem, Alex R. Cook, and Mark Jit. Projecting social contact matrices in 152 countries using contact surveys and demographic data. PLOS Computational Biology, 13(9):e1005697, September 2017. ISSN 1553-7358. doi: 10.1371/journal.pcbi.1005697. URL https://journals.plos.org/ploscompbiol/article?id=10.1371/journal.pcbi.1005697.

[4] Weilin Zhou and Wei Wang. Fast-spreading SARS-CoV-2 variants: challenges to and new design strategies of COVID-19 vaccines. Signal Transduction and Targeted Therapy, 6(1):226, June 2021. ISSN 2059-3635. doi: 10.1038/s41392-021-00644-x. URL https://www.nature.com/articles/s41392-021-00644-x.

[5] Andria Mousa, Peter Winskill, Oliver John Watson, Oliver Ratmann, Mélo die Monod, Marco Ajelli, Aldiouma Diallo, Peter J Dodd, Carlos G Grijalva, Moses Chapa Kiti, Anand Krishnan, Rakesh Kumar, Supriya Kumar, Kin O Kwok, Claudio F Lanata, Olivier Le Polain de Waroux, Kathy Leung, Wiriya Mahikul, Alessia Melegaro, Carl D Morrow, Joël Mossong, Eleanor FG Neal, D James Nokes, Wirichada Panngum, Gail E Potter, Fiona M Russell, Siddhartha Saha, Jonathan D Sugimoto, Wan In Wei, Robin R Wood, Joseph Wu, Juanjuan Zhang, Patrick Walker, and Charles Whittaker. Social contact patterns and implications for infectious disease transmission – a systematic review and meta-analysis of contact surveys. eLife, 10:e70294, November 2021. ISSN 2050-084X. doi: 10.7554/eLife.70294. URL 10.7554/eLife.70294.

[6] Casey F. Breen, Ayesha S. Mahmud, and Dennis M. Feehan. Novel estimates re-veal subnational heterogeneities in disease-relevant contact patterns in the United States. PLOS Computational Biology, 18(12):e1010742, December 2022. ISSN 1553-7358. doi: 10.1371/journal.pcbi.1010742. URL https://journals.plos.org/ploscompbiol/article?id=10.1371/journal.pcbi.1010742.

[7] Cevat Giray Aksoy, Jose Maria Barrero, Nicholas Bloom, Steven J. Davis, Mathias Dolls, and Pablo Zarate. The global persistence of work from home. Proceedings of the National Academy of Sciences, 122(27):e2509892122, July 2025. doi: 10.1073/pnas.2509892122. URL https://www.pnas.org/doi/10.1073/pnas.2509892122.

[8] Christopher I. Jarvis, Pietro Coletti, Jantien A. Backer, James D. Munday, Christel Faes, Philippe Beutels, Christian L. Althaus, Nicola Low, Jacco Wallinga, Niel Hens, and W. John Edmunds. Social contact patterns following the COVID-19 pandemic: a snapshot of post-pandemic behaviour from the CoMix study. Epidemics, 48:100778, September 2024. ISSN 1755-4365. doi: 10.1016/j.epidem.2024.100778. URL https://www.sciencedirect.com/science/article/pii/S1755436524000392.

[9] Lucy Goodfellow, Billy J. Quilty, Kevin van Zandvoort, and W. John Edmunds. Social contact patterns in the United Kingdom following the COVID-19 pandemic: The Reconnect cross-sectional survey. PLOS Medicine, 23(5):e1005038, May 2026. ISSN 1549-1277. doi: 10.1371/journal.pmed.1005038. URL https://pmc.ncbi.nlm.nih.gov/articles/PMC13166901/.

[10] Christopher I. Jarvis, Kevin Van Zandvoort, Amy Gimma, Kiesha Prem, Megan Auzenbergs, Kathleen O’Reilly, Graham Medley, Jon C. Emery, Rein M. G. J. Houben, Nicholas Davies, Emily S. Nightingale, Stefan Flasche, Thibaut Jombart, Joel Hellewell, Sam Abbott, James D. Munday, Nikos I. Bosse, Sebastian Funk, Fiona Sun, Akira Endo, Alicia Rosello, Simon R. Procter, Adam J. Kucharski, Timothy W. Russell, Gwen Knight, Hamish Gibbs, Quentin Leclerc, Billy J. Quilty, Charlie Diamond, Yang Liu, Mark Jit, Samuel Clifford, Carl A. B. Pearson, Rosalind M. Eggo, Arminder K. Deol, Petra Klepac, G. James Rubin, W. John Edmunds, and CMMID COVID-19 working group. Quantifying the impact of physical distance measures on the transmission of COVID-19 in the UK. BMC Medicine, 18(1):124, May 2020. ISSN 1741-7015. doi: 10.1186/s12916-020-01597-8. URL 10.1186/s12916-020-01597-8.

[11] Amy Gimma, James D. Munday, Kerry L. M. Wong, Pietro Coletti, Kevin van Zandvoort, Kiesha Prem, CMMID COVID-19 working Group, Petra Klepac, G. James Ru-bin, Sebastian Funk, W. John Edmunds, and Christopher I. Jarvis. Changes in social contacts in England during the COVID-19 pandemic between March 2020 and March 2021 as measured by the CoMix survey: A repeated cross-sectional study. PLOS Medicine, 19(3):e1003907, March 2022. ISSN 1549-1676. doi: 10.1371/journal.pmed.1003907. URL https://journals.plos.org/plosmedicine/article?id=10.1371/journal.pmed.1003907.

[12] Michele Tizzani, Alessandro De Gaetano, Christopher I. Jarvis, Amy Gimma, Kerry Wong, W. John Edmunds, Philippe Beutels, Niel Hens, Pietro Coletti, and Daniela Paolotti. Impact of tiered measures on social contact and mixing patterns of in Italy during the second wave of COVID-19. BMC Public Health, 23(1):906, May 2023. ISSN 1471-2458. doi: 10.1186/s12889-023-15846-x. URL 10.1186/s12889-023-15846-x.

[13] Maria Litvinova, Shelly Sinclair, Allisandra G. Kummer, Paulo C. Ventura, Trevor Foster, Kayoko Shioda, M. Elizabeth Halloran, Alessandro Vespignani, and Marco Ajelli. Epistorm-Mix: Mapping Social Contact Patterns for Respiratory Pathogen Spread in the Post-Pandemic United States, November 2025. URL https://www.medrxiv.org/content/10.1101/2025.11.20.25340662v1. ISSN: 3067-2007 Pages: 2025.11.20.25340662.

14. US Census Bureau. American Community Survey 1-Year Data 2023. URL https://www.census.gov/data/developers/data-sets/acs-1year/2023.html. Section: Government.

15. Insight Net. Social Contact Surveys – Insight Net, June 2026. URL https://insightnet.us/social-contact-surveys/.

[16] Audrey M. Dorélien, Narmada Venkateswaran, Jiuchen Deng, Kelly Searle, Eva Enns, Giovann Alarcon Espinoza, and Shalini Kulasingam. Quantifying social contact patterns in Minnesota during stay-at-home social distancing order. BMC Infectious Diseases, 23 (1):324, May 2023. ISSN 1471-2334. doi: 10.1186/s12879-022-07968-1. URL 10.1186/s12879-022-07968-1.

[17] Guillaume Béraud, Sabine Kazmercziak, Philippe Beutels, Daniel Levy-Bruhl, Xavier Lenne, Nathalie Mielcarek, Yazdan Yazdanpanah, Pierre-Yves Boëlle, Niel Hens, and Benoit Dervaux. The French Connection: The First Large Population-Based Contact Survey in France Relevant for the Spread of Infectious Diseases. PLoS ONE, 10(7), July 2015. ISSN 1932-6203. doi: 10.1371/journal.pone.0133203. URL https://www.ncbi.nlm.nih.gov/pmc/articles/PMC4503306/.

[18] Roy M. Anderson and Robert M. May. Infectious Diseases of Humans: Dynamics and Control. OUP Oxford, 1991. ISBN 978-0-19-854040-3.

19. Petra Klepac, Adam J Kucharski, Andrew JK Conlan, Stephen Kissler, Maria Tang, Hannah Fry, and Julia R Gog. Contacts in context: large-scale setting-specific social mixing matrices from the BBC Pandemic project. preprint, Epidemiology, February 2020. URL http://medrxiv.org/lookup/doi/10.1101/2020.02.16.20023754.

[20] Dennis M. Feehan and Ayesha S. Mahmud. Quantifying population contact patterns in the United States during the COVID-19 pandemic. Nature Communications, 12(1):893, February 2021. ISSN 2041-1723. doi: 10.1038/s41467-021-20990-2. URL https://www.nature.com/articles/s41467-021-20990-2.

