## Appendix for "Beyond National Averages: Social Contact Patterns in the Upper Midwest"

### Appendix A: Midwest Analytics & Disease Modeling Center Social Contact Survey

**Quota Description**

Target Population

- The survey sample will target the following population: General population age 0+, residents of MN plus residents of nearby states: North Dakota, South Dakota, Iowa, Wisconsin
- English-language survey-takers.
- Eligibility: One hundred percent (100%)
- Screening: Confirm residing in eligible states

Study 1 n=1000 age 0 and above: Non-probability based opt in panels, 600 from MN and 400 from neighboring states

Study 2 n=1000 age 0 and above: Non-probability based opt in panels, 600 from MN and 400 from neighboring states

- 0-12 year olds - parents will complete the survey
- 13-17 year olds - parents will provide consent and pass the survey to teens for independent completion
- 18+ year olds- complete the survey on their own

**Main Questionnaire (including screener, if applicable)**

*Programming Notes:*

- *Code all refusals as -1.*
- *Please make sure all the randomization and rotation are independent*
- *Remove default instructions.*
- *Show instruction for all MP questions “Select all that apply.”*
- *Remove progress bar*
- *Do not prompt on questions unless specify.*
- *All questions are mandatory for opt-in sample*

Base: all respondents	DOV_OFFPANEL [S]:

1. KP
2. OPT-IN

Complete (status = 3): the panelist completes the survey successfully. [https://amp.ipsosinteractive.com/resume.aspx?st=3&uid=[UI](https://amp.ipsosinteractive.com/resume.aspx?st=3&uid)D]&soval=exZe4HczhSOEkY

Screen out (status = 82): the panelist does not qualify for the objective and target of study. [https://amp.ipsosinteractive.com/resume.aspx?st=82&uid=[UI](https://amp.ipsosinteractive.com/resume.aspx?st=82&uid)D]&soval=exZe4HczhSOEkY

Quota-full (status = 83): the panelist does not qualify for the profile. [https://amp.ipsosinteractive.com/resume.aspx?st=83&uid=[UI](https://amp.ipsosinteractive.com/resume.aspx?st=83&uid)D]&soval=exZe4HczhSOEkY

Base: If DOV_OFFPANEL=2	GROUPCODE [S]:

1. PARENTS OF CHILDREN 0-17
2. GEN POP 18+

8

**PART 1**

Base: GROUPCODE=1

**S1. [S]**

Are you the parent or legal guardian of at least one child age 0 through 17 who lives in your household?

1. Yes
2. No

*Terminate if S1 = “No” or refused*

Base: If S1=1

**S2. [N]**

For how many of the children aged 0 to 17 in your household are you the parent or legal guardian?

**[Number box, range 0-10]**

*Terminate if S2 = 0 or refused*

Base: If S2 >= 1

**S3. [Grid with dropdown with range 0-17 for age, and dropdown for sex (“Male”/”Female”)]**

Thank you. In the table below, please enter the age and sex for each of the children age 0 through 17 in your household for whom you are the parent or legal guardian.

*Show as many lines as children listed in S2*

|  | Age **[dropdown]** | Sex **[dropdown]** |
| --- | --- | --- |
| Child 1 | **[range 0-17]** | 1. Male 2. Female |
| … |  |  |

*Terminate if all S3 Age or Sex is refused after prompt.*

*Child Selection Point*

- *Pick one child at random from S3 with no missing age/sex*

*CHILD_AGE [DATA-ONLY]*

*[Record the selected child’s age from S3: 0-17]*

*CHILD_GENDER [DATA-ONLY]*

*[Record the selected child’s gender from S3: 1=Male, 2=Female]*

*CHILD_NAME [DATA-ONLY]*

Your/your x-year-old child

*CHILD_NAME2 [DATA-ONLY]*

9

my x-year-old child

NChild [*DATA-ONLY*]

*[Record the number of eligible children in S3]*

**CPSRACE_CHILD [M]**

Base: If GROUPCODE=1

What is **[**CHILD_NAME**]**’s race and/or ethnicity? *Please select ALL that apply.*

1. White
2. Black or African American
3. Hispanic or Latino
4. Asian
5. A different race

Base: IF CHILD_AGE=13-17

Terminate if PARENT_ CONSENT=2,refused

**PARENT_ CONSENT [S]**

**[**CHILD_NAME**]** is invited to participate in the Midwest Analytics & Disease Modeling Center Social Contact Survey. The survey is being run by Ipsos on behalf of the University of Minnesota and the Minnesota Department of Health. The research is part of a larger project funded by the U.S. Centers for Disease Control and Prevention (CDC) in order to better prepare for disease outbreaks and pandemics.

We will ask how many people **[**CHILD_NAME**]** had contact with **yesterday** at home, school, work, and during leisure time.

Their input is very important for understanding how people, including children interact with each other. **[**CHILD_NAME**]**’s participation in this study is voluntary. They can stop taking the survey at any time. This survey takes about 15 minutes.

The information they give will be kept confidential. The study will not be used for marketing.

I give permission for my child to complete this survey.

1. Yes
2. No

Base: IF CHILD_AGE=0-12

**PARENT_ CONSENT2 [S]**

You are invited to participate in the Midwest Analytics & Disease Modeling Center Social Contact Survey on behalf of [CHILD NAME]. The survey is being run by Ipsos on behalf of the University of Minnesota and the Minnesota Department of Health. The research is part of a larger project funded by the U.S. Centers for Disease Control and Prevention (CDC) in order to better prepare for disease outbreaks and pandemics.

10

We will ask how many people **[**CHILD_NAME**]** had contact with **yesterday** at home, school, work, and during leisure time.

Your input is very important for understanding how people, including children interact with each other. Your participation in this study is voluntary. You can stop taking the survey at any time. This survey takes about 15 minutes.

The information you give will be kept confidential. The study will not be used for marketing.

Base: IF GROUPCODE=2

**INTRO0 [DISP]**

Thank you for agreeing to participate in the Midwest Analytics & Disease Modeling Center Social Contact Survey. The survey is being run by Ipsos on behalf of the University of Minnesota and the Minnesota Department of Health. The research is part of a larger project funded by the U.S. Centers for Disease Control and Prevention (CDC) in order to better prepare for disease outbreaks and pandemics.

We will ask how many people you had contact with **yesterday** at home, school, work, and during leisure time.

Your input is very important for understanding how people interact with each other. Your participation in this study is voluntary. You can stop taking the survey at any time. This survey takes about 15 minutes.

The information you give will be kept confidential. The study will not be used for marketing.

Base: If DOV_OFFPANEL=2

**[PPAGE]**

AGECONS [Q]

**[If CHILD_AGE=13-17:** Before you pass the survey to your child, we have a few last questions for you. Please keep in mind that your answers are confidential and your personal information will also be kept private.**]**

How old are you?

*Type in your age.*

*SCRIPTER: min.=0, max.=120. Show label to right of box: years old. Prompt following nonresponse. Terminate if AGECONS<18.*

Base: If DOV_OFFPANEL=2

**[PPSEX]**

QSEX [S]

11

Are you…?

1. Male
2. Female

Base: If DOV_OFFPANEL=2	CPSRACE [M]

What is your race and/or ethnicity? *Select all answers that apply.*

1. White
2. Black or African American
3. Hispanic or Latino
4. Asian
5. A different race

Base: If DOV_OFFPANEL=2

Terminate if QSTATE NE 35, 41, 42, 44, 45

**[PPSTATEN] [PPREG4]** QSTATE [S]

What is your state of residence?

63. Alabama

94. Alaska

86. Arizona

71. Arkansas

93. California

84. Colorado

16. Connecticut

51. Delaware

53. District of Columbia

59. Florida

58. Georgia

95. Hawaii

82. Idaho

33. Illinois

32. Indiana

42. Iowa

47. Kansas

61. Kentucky

72. Louisiana
11. Maine

52. Maryland

14. Massachusetts

34. Michigan

41. Minnesota

64. Mississippi

43. Missouri

12

81. Montana

46. Nebraska

88. Nevada

12. New Hampshire

22. New Jersey

85. New Mexico

21. New York

56. North Carolina

44. North Dakota

31. Ohio

73. Oklahoma

92. Oregon

23. Pennsylvania

15. Rhode Island

57. South Carolina

45. South Dakota

62. Tennessee

74. Texas

87. Utah

13. Vermont
54. Virginia

91. Washington

55. West Virginia

35. Wisconsin

83. Wyoming

*SCRIPTER: Assign numeric codes per list above, but show full state name in alphabetic order in programmed survey. Show as drop down of all states and Washington DC.*

*Create data-only variables.*

**Variable name:** PPREG4

**Type:** SP

**Variable Text:** Region 4 – based on State of residence

**Response list:**

1. Northeast
2. Midwest
3. South
4. West

| **PPSTATEN** | **PPREG4** |
| --- | --- |
| 11-23 | 1 |
| 31-47 | 2 |
| 51-74 | 3 |
| 81-95 | 4 |

Base: If DOV_OFFPANEL=2

**[PPMSACAT]**

QZIP [Q]

13

What is your zip code?

*SCRIPTER: min.=00000, max.=99999; require a 5-digit response.*

**Variable name:** QZIP

**Type:** SP

**[USE CROSSWALK TABLE BASED ON ZIP_LEVEL_TABLE. XLSX] [PROGRAMMER: TERMINATE IF QZIP DOES NOT MATCH QSTATE]**

Base: If DOV_OFFPANEL=2	QEMPLOY [Q]

How many hours do you usually work for pay or profit per week? Please include hours you work for pay or profit at all your jobs if you have more than one job. If none, enter “0”. If less than an hour in a week, enter “1”.

*SCRIPTER: min.=0, max.=168. Show label to right of box: Hours per week. Do not allow decimals. Prompt following nonresponse. Create data only variable.*

IF QEMPLOY ≥ 35 PPEMPLOY = 1.

IF QEMPLOY ≤ 34 AND QEMPLOY ≥ 1 PPEMPLOY = 2.

IF QEMPLOY = 0 PPEMPLOY = 3.

**Variable name:** PPEMPLOY [S]

**Variable Text:** Current employment status

**Response list:**

1. Working full-time
2. Working part-time
3. Not working

Base: If QEMPLOY>0	QEMP1 [S]

Which of the following best describes your employer?

1. Government
2. Private-for-profit company
3. Non-profit organization including tax exempt and charitable organizations
4. Self-employed
5. Working in the family business

Base: If QEMPLOY>0

**QEMP2 [S]**

In your current (MAIN) job, what kind of work do you do?

1. Management
2. Business Operations (including Marketing)
3. Financial Operations or Financial Services (including Financial Advisor, Broker)

14

4. Computer and Mathematical (including Information Technology)
5. Architecture and Engineering
6. Life, Physical, and Social Sciences
7. Community and Social Services
8. Legal
9. Education, Training, and Library
10. Arts, Design, Entertainment, Sports, and Media
11. Health Diagnosing or Treating Practitioner (such as physician, nurse, dentist, veterinarian, pharmacist)
12. Health Technologist or Technician (such as paramedic, lab technician, dental hygienist)
13. Health Care Support (such as nursing aide, orderly, dental assistant)
14. Protective Service (such as firefighter, law enforcement worker)
15. Food Preparation and Serving
16. Building and Grounds Cleaning and Maintenance
17. Personal Care and Service
18. Sales
19. Office and Administrative Support
20. Farming, Forestry, and Fishing
21. Construction and Extraction
22. Installation, Maintenance, and Repair
23. Precision Production (such as machinist, welder, baker, printer, tailor)
24. Transportation and Material Moving
25. Armed Forces
26. Other, please specify **[O]**

Base: If DOV_OFFPANEL=2	QSCHOOL [S]

Do you currently attend in-person school?

1. Yes
2. No, not in school
3. No, I attend online school/home school

Base: If DOV_OFFPANEL=2

**[PPEDUC] [PPEDUCAT] [PPEDUC5]** QEDUC [S]

What is the highest level of school you have completed?

*Select one answer only.*

1. Some high school or less – no diploma or GED
2. High school graduate – high school diploma or the equivalent (GED)
3. Some college, no degree
4. Associate degree
5. Bachelor’s degree
6. Master’s degree

15

7. Professional or Doctorate degree

Base: If DOV_OFFPANEL=2

**[PPINCIMP]**

QINC [S]

How much is the combined income of all members of YOUR HOUSEHOLD for the PAST 12 MONTHS?

**[SPACE]**

Please include your income PLUS the income of all members living in your household (including cohabiting partners and armed forces members living at home). Please count income BEFORE TAXES and from all sources (such as wages, salaries, tips, net income from a business, interest, dividends, child support, alimony, and Social Security, public assistance, pensions, or retirement benefits).

*Select one answer only.*

1. Below $50,000
2. $50,000 or more
3. Don’t know

*SCRIPTER: Prompt once if question is skipped. Do not show ‘Don’t know’ initially. Show ‘Don’t know’ only*

*with the prompt if question is skipped initially.*

**[PROMPT]**

Your answer will help represent the entire U.S. population and will be kept confidential. Thank you!

Base: If QINC=1	QINC2 [S]

We would like to get a better estimate of your total HOUSEHOLD income in the past 12 months before taxes. Was it...

**[PROMPT]**

Your answer will help represent the entire U.S. population and will be kept confidential. Thank you!

*Select one answer only.*

1. Less than $5,000
2. $5,000 to $7,499
3. $7,500 to $9,999
4. $10,000 to $12,499
5. $12,500 to $14,999
6. $15,000 to $19,999
7. $20,000 to $24,999
8. $25,000 to $29,999
9. $30,000 to $34,999
10. $35,000 to $39,999
11. $40,000 to $49,999

Base: If QINC=2	QINC3 [S]

16

We would like to get a better estimate of your total HOUSEHOLD income in the past 12 months before taxes. Was it...

**[PROMPT]**

Your answer will help represent the entire U.S. population and will be kept confidential. Thank you!

*Select one answer only.*

3. $50,000 to $59,999
4. $60,000 to $74,999
5. $75,000 to $84,999
6. $85,000 to $99,999
7. $100,000 to $124,999
8. $125,000 to $149,999
9. $150,000 to $174,999
10. $175,000 to $199,999
11. $200,000 to $249,999
12. $250,000 or more

Base: If DOV_OFFPANEL=2

**[PPHOUSE4]**

QHOUSE [S]

What is your housing type?

1. One-family house detached from any other house
2. One-family house attached to one or more houses (such as a condo or townhouse)
3. Building with 2 or more apartments
4. Mobile home
5. Boat, RV, van, etc.

Base: If DOV_OFFPANEL=2

**[PPMARIT5]**

QMARIT [S]

Are you currently...?

1. Married
2. Widowed
3. Divorced
4. Separated
5. Never married

**[PROGRAMMER: IF CHILD_AGE=13-17, ASK PART 4 (Intro4 – Q13) HERE BEFORE S5; ELSE FOLLOW THE ORDER]**

Base: IF CHILD_AGE=13-17

**S5 [S]**

Thank you for your participation. Is **[**CHILD_NAME**]** currently available to take the survey?

1. Yes
2. No, but will be available at a later time

Base: IF S5=1

**S5A [DISPLAY]**

Please have **[**CHILD_NAME**]** come to take the survey now. Please remember we’d like to hear their unique opinions about each question as much as possible**.** Please allow them to have privacy when completing the survey.

Base: IF S5=2 OR REFUSED

**S5B [DISPLAY, remove >> button from screen]**

Please have **[**CHILD_NAME**]** take the survey as soon as possible. They can access the survey through your Member Page. Please allow them to have privacy when completing the survey.

**SCRIPTING: [FROM THIS POINT FORWARD IF RESPONDENT BREAKS OFF THEN SET DATACOLLECTIONSTATUS=TIMED OUT AND SURVEY SHOULD RESUME AT S5 SCREEN]**

Base: IF S5=1

**S5C [DISPLAY]**

**Please give them privacy to take this survey. It is important that they know all survey responses are private and confidential.**

The remaining questions are intended for **[**CHILD_NAME**].**

Base: IF CHILD_AGE=13-17

**INTRO0_1 [DISP]**

Thank you for agreeing to participate in the Midwest Analytics & Disease Modeling Center Social Contact Survey. The survey is being run by Ipsos on behalf of the University of Minnesota and the Minnesota Department of Health. The research is part of a larger project funded by the U.S. Centers for Disease Control and Prevention (CDC) in order to better prepare for disease outbreaks and pandemics.

We will ask how many people you had contact with **yesterday** at home, school, work, and during leisure time.

Your input is very important for understanding how people interact with each other. Your participation in this study is voluntary. You can stop taking the survey at any time. This survey takes about 15 minutes.

The information you give will be kept confidential. The study will not be used for marketing.

Base: IF CHILD_AGE=13-17	TEEN_EMPLOY [Q]

How many hours do you usually work for pay or profit per week? Please include hours you work for pay or profit at all your jobs if you have more than one job. If none, enter “0”. If less than an hour in a week, enter “1”.

*SCRIPTER: min.=0, max.=168. Show label to right of box: Hours per week. Do not allow decimals. Prompt following nonresponse.*

Base: If TEEN_EMPLOY>0	TEEN_QEMP1 [S]

Which of the following best describes your employer?

1. Government
2. Private-for-profit company
3. Non-profit organization including tax exempt and charitable organizations
4. Self-employed
5. Working in the family business

Base: If TEEN_EMPLOY>0

**TEEN_QEMP2 [S]**

In your current (MAIN) job, what kind of work do you do?

1. Management
2. Business Operations (including Marketing)
3. Financial Operations or Financial Services (including Financial Advisor, Broker)
4. Computer and Mathematical (including Information Technology)
5. Architecture and Engineering
6. Life, Physical, and Social Sciences
7. Community and Social Services
8. Legal
9. Education, Training, and Library
10. Arts, Design, Entertainment, Sports, and Media
11. Health Diagnosing or Treating Practitioner (such as physician, nurse, dentist, veterinarian, pharmacist)
12. Health Technologist or Technician (such as paramedic, lab technician, dental hygienist)
13. Health Care Support (such as nursing aide, orderly, dental assistant)
14. Protective Service (such as firefighter, law enforcement worker)
15. Food Preparation and Serving
16. Building and Grounds Cleaning and Maintenance
17. Personal Care and Service
18. Sales
19. Office and Administrative Support
20. Farming, Forestry, and Fishing
21. Construction and Extraction
22. Installation, Maintenance, and Repair
23. Precision Production (such as machinist, welder, baker, printer, tailor)
24. Transportation and Material Moving
25. Armed Forces
26. Other, please specify **[O]**

19

Base: IF GROUPCODE=1

**TEEN_SCHOOL [S]**

**[CHILD_AGE=13-17]**Do you currently attend in-person school, after school care, camp, or other childcare setting?

**[CHILD_AGE=0-12]**Does **[**CHILD_NAME**]** currently attend in-person school, daycare, camp, or any other childcare setting?

1. Yes
2. No, not in school
3. No, currently attending online school/home school

**PART 2 Household contacts**

Base: all respondents

**INTRO1 [DISP]**

**[IF GROUPCODE=2 or CHILD_AGE=13-17:** First, we would like to ask about contacts with people in your **household**, including people who live in the same home as you, such as family members or roommates who share a common area like a kitchen or a bathroom.**]**

**[IF CHILD_AGE=0-12:** First, we would like to ask about contacts with people in your **household**, including people who live in the same home as **[**CHILD_NAME**]**, such as family members or roommates who share a common area like a kitchen or a bathroom.**]**

Base: all respondents

**Q2ZZ1 [N]**

**[IF GROUPCODE=2 or CHILD_AGE=13-17:**How many adults aged 18 years or older usually live or stay in your home with you? Do **NOT** include yourself.**]**

**[IF CHILD_AGE=0-12:** How many adults aged 18 years or older usually live or stay in your home?**. Include yourself in this count.]**

**[N, RANGE: 0-10]** number of adults aged 18 or older

**[IF GROUPCODE=2 or CHILD_AGE=13-17]**How many children aged 17 years or younger usually live or stay in your home with you? Do **NOT** include yourself.

**[IF CHILD_AGE=0-12]**How many children aged 17 years or younger usually live or stay in your home? **Do NOT include [CHILD_NAME].**

**[N, RANGE: 0-10]** number of children aged 17 or younger

Base: If Q2ZZ1 total>=1

**Q2ZZ2. [Grid with N range 0-120 for age, prompt once if any age is refused]**

In the table below, please enter the name (such as first name, initials, or nickname) and age for each of the adults and children in your household. **[IF GROUPCODE=2 or CHILD_AGE=13-17: Please exclude yourself; IF CHILD_AGE=0-12: Please exclude [CHILD_NAME], but include yourself.]**

This name is only needed to make it easier for you to complete the survey, so please pick a name that will help you identify each person later in the survey. Names are not visible to anyone outside of this survey.

20

*Show as many lines as listed in Q2ZZ1 total, and then have a button to add 5 rows per click, max 40 rows*

| Name | Age **[N]** |
| --- | --- |
| **[O]** | **[range 0-120]** |
| … |  |

**[PROGRAMMER: loop Q2C-Q2D for each Name entered in Q2ZZ2]**

Base: at least one initial is entered in Q2ZZ2 Put Q2C and Q2D on the same screen

**Q2C. [S]**

What sex is **[**NAME_HH**]**?

1. Male
2. Female

**Q2D. [M]**

What is the race/ethnicity of **[**NAME_HH**]**? *Please select ALL that apply.*

1. White
2. Black or African American
3. Hispanic or Latino
4. Asian
5. A different race

Base: all respondents

**Q2ZZ3 [S]**

**[IF GROUPCODE=2 or CHILD_AGE=13-17:**Do you; **IF CHILD_AGE=0-12:** Does **[**CHILD_NAME**]]** have a

medical condition that puts **[IF GROUPCODE=2 or CHILD_AGE=13-17:** you; **IF CHILD_AGE=0-12:** them**]** at high-risk of getting very sick from an infectious disease?

High-risk conditions can include **[IF GROUPCODE=2 or CHILD_AGE=13-17:** pregnancy or**]** diseases such as diabetes, immunosuppression, obesity, and chronic diseases that affect the lungs, heart, kidney, liver, or nervous system.

1. Yes
2. No
3. I don’t know

Base: If Q2ZZ1 total>=1

**Q2ZZ4 [S]**

Do any of your household members (not including **[IF GROUPCODE=2 or CHILD_AGE=13-17:** yourself; **IF CHILD_AGE=0-12: [**CHILD_NAME**]]**) have a medical condition that puts them at high-risk of getting very sick from an infectious disease?

High-risk conditions can include pregnancy or diseases such as diabetes, immunosuppression, obesity, and chronic diseases that affect the lungs, heart, kidney, liver, or nervous system.

1. Yes
2. No
3. I don’t know

Base: at least one initial is entered in Q2ZZ2

**INTRO 2 [DISP]**

**[IF GROUPCODE=2 or CHILD_AGE=13-17:** We will now ask you to remember which household members you had direct contact with **yesterday, [Month, day, year]**, from the time you woke up at the start of your day until you went to sleep at the end of the day. A contact includes people you had an in-person **conversation** of at least 5 words with, or people you had **physical contact** with (such as a handshake, hug, kiss, or contact sports).**]**

**[IF CHILD_AGE=0-12:** With **[**CHILD_NAME**]**’s help, we will now ask you to remember which household members your child had direct contact with **yesterday, [Month, day, year]**, from the time they woke up at the start of the day until they went to sleep at the end of the day. A contact includes people they had an **in-person conversation** of at least 5 words with, or people they had **physical contact** with (such as a handshake, hug, kiss, or contact sports).**]**

**If [IF GROUPCODE=2 or CHILD_AGE=13-17: you; [IF CHILD_AGE=0-12: they] only spoke to someone over the phone or internet, they should not be included.**

Base: at least one initial is entered in Q2ZZ2

**Q2E [Accordion, S]**

Yesterday, did **[IF GROUPCODE=2 or CHILD_AGE=13-17:** you; **IF CHILD_AGE=0-12: [**CHILD_NAME**]]**

interact with…?

Item:

**[LIST NAME_HH from Q2ZZ2]**

Scale:

1. No, **[IF GROUPCODE=2 or CHILD_AGE=13-17:** I; **IF CHILD_AGE=0-12: [**CHILD_NAME2**]]** did not

have an in-person conversation or any physical contact with them

2. Yes, **[IF GROUPCODE=2 or CHILD_AGE=13-17:** I; **IF CHILD_AGE=0-12: [**CHILD_NAME2**]]** had an

in-person conversation of at least 5 words

3. Yes, **[IF GROUPCODE=2 or CHILD_AGE=13-17:** I; **IF CHILD_AGE=0-12: [**CHILD_NAME2**]]** had

physical contact (such as a hand shake, hug, or kiss)

4. Both (in-person conversation and physical contact)

**[PROGRAMMER: loop Q2G-Q2F for each Name if Q2E=2,3,4]**

Base: If Q2E=2,3,4 for listed NAME

**Q2G [M]**

Next, we would like to know more about **[IF GROUPCODE=2 or CHILD_AGE=13-17:** your; **IF CHILD_AGE=0-12:** their**]** interaction with **[**NAME_HH**]** yesterday.

Where did **[IF CHILD_AGE=13-17 or GROUPCODE=2:**you; **IF CHILD_AGE=0-12:[**CHILD_NAME**]]** interact

with **[**NAME_HH**]**? *Please select ALL that apply.*

1. Your home (including at your door, in your garden, and within entrances to your home, such as stairways, elevators, and hallways)
2. Your workplace
3. **[IF CHILD_AGE=13-17 or GROUPCODE=2:**School, pre-school; **IF CHILD_AGE=0-12:**Their school**]**, daycare, home-based daycare, afterschool care, or summer camp
4. Someone else’s house (including at their door, garden, and within entrances of their home, such as stairways, elevators, and hallways)
5. A place of worship
6. On public transport
7. At a supermarket, grocery store, or retail pharmacy
8. At another type of shop
9. At a place of entertainment, such as a restaurant, bar, or cinema
10. At a place for indoor sports, such as a gym or stadium (including outdoor stadiums with indoor areas)
11. At a healthcare facility, such as a hospital or clinic
12. Outside, such as in a park, on the street, or an outdoor sporting event
13. Somewhere else, please specify: **[O]**

Base: If Q2E=2,3,4 for listed NAME

**Q2H [S]**

In total, about how much time did **[IF GROUPCODE=2 or CHILD_AGE=13-17:** you; **IF CHILD_AGE=0-12: [**CHILD_NAME**]]** spend with **[**NAME_HH**]** in person yesterday?

1. Less than 5 minutes
2. 5 to less than 15 minutes
3. 15 minutes to less than 1 hour
4. 1 to 4 hours
5. More than 4 hours

Base: If Q2E=2,3,4 for listed NAME

**Q2F [S]**

What is **[**NAME_HH**]**’s relationship to **[IF GROUPCODE=2 or CHILD_AGE=13-17:** you; **IF CHILD_AGE=0-12: [**CHILD_NAME**]]**?

1. **[IF GROUPCODE=2 or CHILD_AGE=13-17]**My spouse or romantic partner
2. **[IF GROUPCODE=2 or CHILD_AGE=13-17:** My; **IF CHILD_AGE=0-12:** Child’s**]** parent
3. **[IF GROUPCODE=2 or CHILD_AGE=13-17]**My child
4. **[IF GROUPCODE=2 or CHILD_AGE=13-17:** My; **IF CHILD_AGE=0-12:** Child’s**]** sibling
5. **[IF GROUPCODE=2 or CHILD_AGE=13-17:** My; **IF CHILD_AGE=0-12:** Child’s**]** grandparent
6. **[IF GROUPCODE=2 or CHILD_AGE=13-17:** Other; **IF CHILD_AGE=0-12:** Child’s other**]** relative
7. Housemate, not a relative

**PART 3 Non-household member contacts**

Base: all respondents

**INTRO3 [DISP]**

**[IF GROUPCODE=2 or CHILD_AGE=13-17:** Next, we would like to know about people **who live outside of your household** who you interacted with **yesterday, [Month, day, year]**, from the time you wok

e up at

23

the start of your day until you went to sleep at the end of the day. This could include friends, family, work colleagues, people you spoke to in shops, and so on. As a reminder, direct contacts include people you had an **in-person conversation** of at least 5 words with, or people you had **physical contact** with (such as a handshake, hug, kiss, or contact sports). If you have a large number of contacts, **we will only ask about details for a subset of your contacts.]**

**[IF CHILD_AGE=0-12:** Next, we would like to know about people **who live outside of your household** who **[**CHILD_NAME**]** interacted with **yesterday, [Month, day, year]**, from the time they woke up at the start of the day until they went to sleep at the end of the day. This could include friends, family, people they spoke to in shops, and so on. As a reminder, direct contacts include people they had an **in-person conversation** of at least 5 words with, or people they had **physical contact** with (such as a handshake, hug, kiss, or contact sports). If **[**CHILD_NAME**]** has a large number of contacts, **we will only ask about details for a subset of their contacts.]**

If **[IF GROUPCODE=2 or CHILD_AGE=13-17:** you; **IF CHILD_AGE=0-12:** they**]** had multiple contacts with the same person **yesterday**, only count the contact once.

**PART 3A Work contacts**

Base: IF (CHILD_AGE=13-17 and TEEN_EMPLOY>0) or (GROUPCODE=2 and QEMPLOY>0)

**Q3A [N]**

**At work yesterday**, with how many people did you have an **in-person** conversation of at least 5 words or physical contact?

Do **NOT** count people that you also saw outside of work (for example, a spouse or partner, or a colleague you went to dinner with), these contacts will be asked about in the next section.

Number of contacts: **[N, range 0-200]**

[S] I was not at work yesterday. [SKIP to Part 3B]

Base: Q3A>0

**Q3A_1 [N]**

In the table below, please enter the name (such as first name, initials, or nickname) of **people you only interacted with at work** that you talked to **in person** or had **physical contact** with yesterday.

This name is only needed to make it easier for you to complete the survey, so please pick a name that will help you identify each person later in the survey. Names are not visible to anyone outside of this survey.

*Show as many lines as listed in Q3A, and then have a button to add 5 rows per click, max 40 rows*

Name

**[O]**

…

**[PROGRAMMER: if N Q3A_1<=10, loop Q3A_5 - Q3A_6 for each WORK Name entered in Q3A_1; ELSE if Q3A_1>10, randomly select 10 names and ask detailed follow up questions]**

24

Base: at least one NAME is entered in Q3A_1 Put Q3A_5 and Q3A_8 on the same screen

**Q3A_5 [S]**

Next, we would like to know more about your interaction with **[**NAME_WORK**]** yesterday.

When you interacted with **[**NAME_WORK**]** yesterday, did you have...?

1. An in-person conversation of at least 5 words
2. Physical contact (such as a handshake, hug, or kiss)
3. Both (in-person conversation and physical contact)

**Q3A_8 [S]**

In total, about how much time did you spend with **[**NAME_WORK**]** in person yesterday?

1. Less than 5 minutes
2. 5 to less than 15 minutes
3. 15 minutes to less than 1 hour
4. 1 to 4 hours
5. More than 4 hours

Base: at least one NAME is entered in Q3A_1 Put Q3A_7 and Q3A_9 on the same screen

**Q3A_7 [S]**

Was your interaction with **[**NAME_WORK**]** yesterday inside or outside?

1. Inside
2. Outside
3. Both

**Q3A_9 [S]**

How often do you usually interact in person with **[**NAME_WORK**]**?

1. Almost every day
2. Once or twice a week
3. Every 2 to 3 weeks
4. Once a month
5. Less than once a month
6. Never met them before

Base: at least one NAME is entered in Q3A_1 Put Q3A_3 and Q3A_4 on the same screen

**Q3A_3 [S]**

What sex is **[**NAME_WORK**]**? Please guess if you are not sure.

1. Male
2. Female

**Q3A_4 [M]**

25

What is the race/ethnicity of **[**NAME_WORK**]**? Please guess if you are not sure. *Please select ALL that apply.*

1. White
2. Black or African American
3. Hispanic or Latino
4. Asian
5. A different race

Base: at least one NAME is entered in Q3A_1 Put Q3A_2 and Q3A_6 on the same screen

**Q3A_2 [N, 0-120]**

What is the age of **[**NAME_WORK**]**? Please give an estimate if you are not sure.

**[N]** years old

**Q3A_6 [S]**

What is **[**NAME_WORK**]**’s relationship to you?

1. My colleague, supervisor, or boss
2. My customer, client, or patient
3. Other, please specify: **[O]**

Base: at least 11 NAME_WORK are entered in Q3A_1

**Q3A_10 [N]**

For the remaining **[**N NAME_WORK-10**]** of your contacts at work yesterday, what age group are they in?

*Total should match N NAME_WORK-5. Prompt if total does not match.*

| **Age Group** | **Number of Contacts** |
| --- | --- |
| Under 1 |  |
| 1-4 |  |
| 5-9 |  |
| 10-17 |  |
| 18-29 |  |
| 30-39 |  |
| 40-49 |  |
| 50-59 |  |

| 60-69 |  |
| --- | --- |
| 70-79 |  |
| 80+ |  |
| TOTAL | **[Auto calculate]** |

**PART 3B School contacts**

Base: IF CHILD_AGE=0-12

**Q3B0 [M]**

Yesterday, did **[**CHILD_NAME**]** attend any of the following? *Please select ALL that apply.*

1. School
2. Pre-school
3. Daycare (including home-based daycare)
4. An afterschool program
5. Summer camp
6. None of the above **[S, SKIP to Q3C]**

Base: IF Q3B0=1,2	Q3B0A [N]

About many students are in **[CHILD_NAME]**’s class?

[N, RANGE 0-999] students

Base: IF (CHILD_AGE=13-17 and TEEN_SCHOOL=1) or (GROUPCODE=2 and QSCHOOL=1) or Q3B0=1,2,3,4,5

**Q3B [N, Range 0-40]**

**[IF CHILD_AGE=13-17 or GROUPCODE=2):**If you are attending classes at a school, college or university, how many people did you have contact with **yesterday** at your school, college or university?

If you work at a school, exclude people already counted at work. Also, do **not** include people that you **also** saw outside of school in this number (for example, a classmate you had dinner with), you will be asked about these contacts in a later section.**]**

**[IF CHILD_AGE=0-12:** When **[**CHILD_NAME**]** attended **[**INSERT RESPONSES FROM Q3B0**]** yesterday, how many people did they have an **in-person conversation** of at least 5 words with or have **physical** contact with in these locations? You can ask your child for input, or your best guest is fine.

If you aren’t sure, think about how many children are in this child’s class(es), and how many teachers or

adults they might interact with during the day as well, to get an approximate number.

Do **not** include people that your child **also** saw outside of school in this number (for example a school friend who came over your house to play), you will be asked about these contacts in a later section.**]**

Number of contacts: **[N, range 0-200]**

**[S] [IF CHILD_AGE=13-17 or GROUPCODE=2):**I was not in school yesterday**; IF CHILD_AGE=0-12:**They were not in school yesterday**]**.

Base: Q3B>0

**Q3B_1 [N]**

Please write the nicknames of people **[IF CHILD_AGE=13-17 or GROUPCODE=2:**you; **IF CHILD_AGE=0-12:[**CHILD_NAME**]] ONLY interacted with at SCHOOL** that **[IF CHILD_AGE=13-17 or GROUPCODE=2:** you**; IF CHILD_AGE=0-12:** they**]** talked to in person or had physical contact with yesterday.

Note that this nickname/initials is only needed to make it easier for you to complete the survey, so please pick a nickname that will help you identify each contact person later in the survey. Nicknames are not visible to anyone outside of this survey.

*Show as many lines as listed in Q3B, and then have a button to add 5 rows per click, max 40 rows*

Name

**[O]**

…

**[PROGRAMMER: if N Q3B_1<=10, loop Q3B_5 - Q3B_6 for each SCHOOL Name entered in Q3B_1; ELSE if Q3B_1>10, randomly select 10 names and ask detailed follow up questions]**

Base: at least one NAME is entered in Q3B_1 Put Q3B_5 and Q3B_8 on the same screen

**Q3B_5 [S]**

**[IF CHILD_AGE=13-17 or GROUPCODE=2:**Next, we would like to know more about your interaction with

**[**NAME_SCHOOL**]** yesterday.

When you had direct contact with **[**NAME_SCHOOL**]** yesterday, to your recollection did you have...?**] [IF CHILD_AGE=0-12:** Next, we would like to know more about your child’s interaction with **[**NAME_SCHOOL**]** yesterday.

When **[**CHILD_NAME**]** had direct contact with **[**NAME_SCHOOL**]** yesterday, did they have...?**]**

1. An in-person conversation of at least 5 words
2. Physical contact (such as a handshake, hug, or kiss)
3. Both (in-person conversation and physical contact)

**Q3B_8 [S]**

In total, about how much time did **[IF CHILD_AGE=13-17 or GROUPCODE=2:**you; **IF CHILD_AGE=0-12:[**CHILD_NAME**]]** spend with **[**NAME_SCHOOL**]** in person yesterday?

1. Less than 5 minutes
2. 5 to less than 15 minutes
3. 15 minutes to less than 1 hour

28

4. 1 to 4 hours

29

5. More than 4 hours

Base: at least one NAME is entered in Q3B_1 Put Q3B_7 and Q3B_9 on the same screen

**Q3B_7 [S]**

**[IF CHILD_AGE=13-17 or GROUPCODE=2:**Was your interaction with **[**NAME_SCHOOL**]** yesterday inside or outside?**]**

**[IF CHILD_AGE=12:** Was the time **[**CHILD_NAME**]** spent with **[**NAME_SCHOOL] yesterday inside or outside?**]**

1. Inside
2. Outside
3. Both

**Q3B_9 [S]**

How often **[IF CHILD_AGE=13-17 or GROUPCODE=2:**do you**; IF CHILD_AGE=0-12:**does **[**CHILD_NAME**]]**

usually interact in person with **[**NAME_SCHOOL**]**?

1. Almost every day
2. Once or twice a week
3. Every 2 to 3 weeks
4. Once a month
5. Less than once a month
6. Never met them before

Base: at least one NAME is entered in Q3B_1 Put Q3B_3 and Q3B_4 on the same screen

**Q3B_3 [S]**

What sex is **[**NAME_SCHOOL**]**? Please guess if you are not sure.

1. Male
2. Female

**Q3B_4 [M]**

What is the race/ethnicity of **[**NAME_SCHOOL**]**? Please guess if you are not sure. *Please select ALL that apply.*

1. White
2. Black or African American
3. Hispanic or Latino
4. Asian
5. A different race

Base: at least one NAME is entered in Q3B_1 Put Q3B_2 and Q3B_6 on the same screen

**Q3B_2 [N, 0-120]**

What is the age of **[**NAME_SCHOOL**]**? Please give an estimate if you are not sure.

**[N]** years old

**Q3B_6 [S]**

What is **[**NAME_SCHOOL**]**’s relationship to **[IF CHILD_AGE=13-17 or GROUPCODE=2:**you**; IF CHILD_AGE=0-12:[**CHILD_NAME**]]**?

1. **[IF CHILD_AGE=13-17 or GROUPCODE=2:**My**; IF CHILD_AGE=0-12:**Their**]** classmate
2. **[IF CHILD_AGE=13-17 or GROUPCODE=2:**My teacher/school employee**; IF CHILD_AGE=0-12:**Their teacher/coach/counselor/school employee**]**
3. Community member
4. Other, please specify: **[O]**

Base: at least 11 NAME_SCHOOL are entered in Q3B_1

**Q3B_10 [N]**

For the remaining **[**N NAME_SCHOOL-10**]** of **[IF CHILD_AGE=13-17 or GROUPCODE=2:**your contacts at school yesterday**; IF CHILD_AGE=0-12:[**CHILD_NAME**]**’s contacts at school**]**, what age group are they in:

*Total should match N NAME_SCHOOL-5. Prompt if total does not match.*

| **Age Group** | **Number of Contacts** |
| --- | --- |
| Under 1 |  |
| 1-4 |  |
| 5-9 |  |
| 10-17 |  |
| 18-29 |  |
| 30-39 |  |
| 40-49 |  |
| 50-59 |  |
| 60-69 |  |
| 70-79 |  |
| 80+ |  |
| TOTAL | **[Auto calculate]** |

**PART 3C Non-household member contacts outside of school and work**

Base: all respondents

**Q3C [N]**

**[IF CHILD_AGE=13-17 or GROUPCODE=2:Yesterday**, **outside of your household members and interactions at work and school**, how many people did you have an **in-person conversation** of at least 5 words with or have **physical contact** with?**]**

**[IF CHILD_AGE=0-12:Yesterday**, outside of **[**CHILD_NAME**]**’s household members and interactions at school, how many people did they have an **in-person conversation** of at least 5 words with or have **physical contac**t with?**]**

Number of contacts: **[N, range 0-200]**

[S] **[IF CHILD_AGE=13-17 or GROUPCODE=2:**None, I did not have in-person interactions with anyone who lives outside of my household, or outside of work or school; **IF CHILD_AGE=0-12:**None, my child did not have in-person interactions with anyone who lives outside of my household, or outside of school**]**

**[calculate total_contacts = work_total (Q3A) + school_total (Q3B) + other_loc_total (Q3C)]**

Base: Q3C>0

**Q3C_1 [O]**

**[IF CHILD_AGE=13-17 or GROUPCODE=2:**Please write the nicknames of people you **ONLY interacted with outside of your household, or outside of work or school** that you talked to in person or had physical contact with yesterday.**]**

**[IF CHILD_AGE=0-12:** Please write the nicknames of people **[**CHILD_NAME**] ONLY interacted with outside of their household, or outside of school** that they talked to in person or had physical contact with yesterday.**]**

*Show as many lines as listed in Q3C, and then have a button to add 5 rows per click, max 40 rows*

Name

**[O]**

…

**[PROGRAMMER: if N Q3C_1<=10, loop Q3C_5 - Q3C_6 for each OTHER Name entered in Q3C_1; ELSE if Q3C_1>10, randomly select 10 names and ask detailed follow up questions]**

Base: at least one NAME is entered in Q3C_1 Put Q3C_5 and Q3C_8 on the same screen

**Q3C_5 [S]**

**[IF CHILD_AGE=13-17 or GROUPCODE=2:** Next, we would like to know more about your interaction with

**[**NAME_OTHER**]** yesterday.

When you interacted with **[**NAME_OTHER**]** yesterday, did you have...?**]**

**[IF CHILD_AGE=0-12:** Next, we would like to know more about your child’s interaction with

**[**NAME_OTHER**]** yesterday.

When **[**CHILD_NAME**]** interacted with **[**NAME_OTHER**]** yesterday, did they have...?**]**

1. An in-person conversation of at least 5 words
2. Physical contact (such as a handshake, hug, or kiss)
3. Both (in-person conversation and physical contact)

**Q3C_8 [S]**

In total, about how much time did **[IF CHILD_AGE=13-17 or GROUPCODE=2:**you; **IF CHILD_AGE=0-12:[**CHILD_NAME**]]** spend with **[**NAME_OTHER**]** in person yesterday?

1. Less than 5 minutes
2. 5 to less than 15 minutes
3. 15 minutes to less than 1 hour
4. 1 to 4 hours
5. More than 4 hours

Base: at least one NAME is entered in Q3C_1

**Q3C_11 [M]**

Where did **[IF CHILD_AGE=13-17 or GROUPCODE=2:**you; **IF CHILD_AGE=0-12:[**CHILD_NAME**]]** interact

with **[**NAME_ OTHER**]**? *Please select ALL that apply.*

14. Your home (including at your door, in your garden, and within entrances to your home, such as stairways, elevators, and hallways)
15. Your workplace
16. **[IF CHILD_AGE=13-17 or GROUPCODE=2:**School, pre-school; **IF CHILD_AGE=0-12:**Their school**]**, daycare, home-based daycare, afterschool care, or summer camp
17. Someone else’s house (including at their door, garden, and within entrances of their home, such as stairways, elevators, and hallways)
18. A place of worship
19. On public transport
20. At a supermarket, grocery store, or retail pharmacy
21. At another type of shop
22. At a place of entertainment, such as a restaurant, bar, or cinema
23. At a place for indoor sports, such as a gym or stadium (including outdoor stadiums with indoor areas)
24. At a healthcare facility, such as a hospital or clinic
25. Outside, such as in a park, on the street, or an outdoor sporting event
26. Somewhere else, please specify: **[O]**

Base: at least one NAME is entered in Q3C_1 Put Q3C_7 and Q3C_9 on the same screen

**Q3C_7 [S]**

Was **[IF CHILD_AGE=13-17 or GROUPCODE=2:**your; **IF CHILD_AGE=0-12:[**CHILD_NAME**]**’s**]** interaction

with **[**NAME_OTHER**]** yesterday inside or outside?

1. Inside
2. Outside
3. Both

**Q3C_9 [S]**

32

How often **[IF CHILD_AGE=13-17 or GROUPCODE=2:**do you; **IF CHILD_AGE=0-12:**does **[**CHILD_NAME**]]**

usually interact in person with **[**NAME_OTHER**]**?

1. Almost every day
2. Once or twice a week
3. Every 2 to 3 weeks
4. Once a month
5. Less than once a month
6. Never met them before

Base: at least one NAME is entered in Q3C_1 Put Q3C_3 and Q3C_4 on the same screen

**Q3C_3 [S]**

What sex is **[**NAME_OTHER**]**? Please guess if you are not sure.

1. Male
2. Female

**Q3C_4 [M]**

What is the race/ethnicity of **[**NAME_OTHER**]**? Please guess if you are not sure. *Please select ALL that apply.*

1. White
2. Black or African American
3. Hispanic or Latino
4. Asian
5. A different race

Base: at least one NAME is entered in Q3C_1 Put Q3C_2 and Q3C_6 on the same screen

**Q3C_2 [N, 0-120]**

What is the age of **[**NAME_ OTHER**]**? Please give an estimate if you are not sure.

**[N]** years old

**Q3C_6 [S]**

What is **[**NAME_ OTHER**]**’s relationship to **[IF CHILD_AGE=13-17 or GROUPCODE=2:**you; **IF CHILD_AGE=0-12:[**CHILD_NAME**]]**?

1. **[IF CHILD_AGE=13-17 or GROUPCODE=2]**My spouse or romantic partner (not living in my household)
2. **[IF CHILD_AGE=13-17 or GROUPCODE=2:**My; **IF CHILD_AGE=0-12:**Their**]** family
3. **[IF CHILD_AGE=13-17 or GROUPCODE=2:**My; **IF CHILD_AGE=0-12:**Their**]** friend
4. **[IF CHILD_AGE=13-17 or GROUPCODE=2:**My work colleague, teacher, or classmate**; IF CHILD_AGE=0-12:**Their teacher, camp counselor, or classmate**]**
5. A neighbor or community member
6. **[IF CHILD_AGE=13-17 or GROUPCODE=2]**I am this person’s customer, client, or patient

33

7. **[IF CHILD_AGE=13-17 or GROUPCODE=2]**This person is my customer, client, or patient
8. Other, please specify: **[O]**

Base: at least 11 NAME_OTHER are entered in Q3C_1

**Q3C_10 [N]**

For the remaining **[**N NAME_OTHER-10**]** of **[IF CHILD_AGE=13-17 or GROUPCODE=2:**your**; IF CHILD_AGE=0-12:[**CHILD_NAME**]**’s**]** contacts that you have not already described, what age group are they in:

*Total should match N NAME_OTHER-5. Prompt if total does not match.*

| **Age Group** | **Number of Contacts** |
| --- | --- |
| Under 1 |  |
| 1-4 |  |
| 5-9 |  |
| 10-17 |  |
| 18-29 |  |
| 30-39 |  |
| 40-49 |  |
| 50-59 |  |
| 60-69 |  |
| 70-79 |  |
| 80+ |  |
| TOTAL | **[Auto calculate]** |

Base: IF Q3C>40

**Q3C_12 [S]**

Where did the majority of these interactions take place? If these interactions took place in more than one location, which is the primary one?

1. Your home (including at your door, in your garden, and within entrances to your home, such as stairways, elevators, and hallways)
2. Your workplace
3. **[IF CHILD_AGE=13-17 or GROUPCODE=2:**School, pre-school; **IF CHILD_AGE=0-12:**Their school**]**, daycare, home-based daycare, afterschool care, or summer camp
4. Someone else’s house (including at their door, garden, and within entrances of their home, such as stairways, elevators, and hallways)

34

5. A place of worship
6. On public transport
7. At a supermarket, grocery store, or retail pharmacy
8. At another type of shop
9. At a place of entertainment, such as a restaurant, bar, or cinema
10. At a place for indoor sports, such as a gym or stadium (including outdoor stadiums with indoor areas)
11. At a healthcare facility, such as a hospital or clinic
12. Outside, such as in a park, on the street, or an outdoor sporting event
13. Somewhere else, please specify: **[O]**

Base: all respondents

**Q6 [S]**

Was yesterday a typical **[if date-1=weekday:** weekday; **if date-1=weekend:** weekend day**]** for **[IF CHILD_AGE=13-17 or GROUPCODE=2:**you; **IF CHILD_AGE=0-12:[**CHILD_NAME**]]**?

1. Yes, it was a typical day and number of interactions
2. No, **[IF CHILD_AGE=13-17 or GROUPCODE=2:**I; **IF CHILD_AGE=0-12:**they**]** typically have **more** in-person interactions
3. No, **[IF CHILD_AGE=13-17 or GROUPCODE=2:**I; **IF CHILD_AGE=0-12:**they**]** typically have **less** in-person interactions

**PART 4 Additional questions**

Base: all respondents

**Intro4 [DISP]**

**[If GROUPCODE=2 OR CHILD_AGE=0-12:**Thank you for taking the time to tell us about **[If GROUPCODE=2:**your**; if CHILD_AGE=0-12:**your child’s**]** in-person interactions yesterday.**]** The remaining questions ask about topics that might be relevant to future public health concerns.

Base: IF QEMPLOY>0

**Q7 [S]**

Where do you work?

1. 100% outside of the home (ex. at an office or workplace)
2. 100% at home
3. Mix of home and workplace
4. Another location that is not my home or workplace, such as a coffee shop or library

Base: Q7=3

**Q7b [S]**

Do you work ….?

1. Mostly home
2. Mostly at workplace
3. Equally between home and work

35

Base: IF QEMPLOY>0

**Q8 [S]**

Could you do your job from home if needed?

1. Definitely yes
2. Probably yes
3. Probably not
4. Definitely not

Base: all respondents

**Q9A [S]**

Do you currently live or work on a farm?

1. Yes
2. No

Base: all respondents

**Q9B [S]**

Do you own any livestock, such as backyard chickens, horses? Do NOT include cats and dogs.

1. Yes
2. No

Base: If Q9A=1 OR Q9B=1

**Q10 [M]**

On a typical day, do you have any direct contact with the following animals? *Select all that apply.*

1. Pigs
2. Cows
3. Poultry, such as chickens or turkeys
4. Waterfowl, such as ducks, geese, or swans
5. Other animals
6. None of these **[S]**

Base: all respondents

**Q11 [Accordion, S]**

If public health officials recommended wearing a mask (similar to the early COVID-19 pandemic), would you choose to wear a mask in the following settings if you visited them tomorrow?

Item:

1. While visiting with friends or family at your home or their home (including at the door, in the garden, and within entrances to the home such as stairways, elevators, and hallways)
2. Your workplace
3. School, pre-school, daycare, home-based daycare, afterschool care, or summer camp
4. A place of worship
5. On public transport

36

6. At an airport or on an airplane
7. At a supermarket, grocery store, or retail pharmacy
8. **Inside** another type of shop
9. At an **outside** shop or vendor, such as a food truck or farmers market
10. At a place for indoor sports, such as a gym or stadium (including outdoor stadiums with indoor areas)
11. At a healthcare facility, such as a hospital or clinic
12. Outside, such as in a park, on the street, or outdoor sporting event

Scale:

1. Yes, I would wear a mask regardless of recommendations or requirements
2. Yes, but only if it was recommended by health officials
3. Yes, but only if it was required
4. No, I would not wear a mask even if it was recommended or required

Base: all respondents

**Q12 [S]**

If you had to stop interacting with people in person due to a disease outbreak, would you be able to reduce the number of people you interact with outside of your household?

1. I could reduce my non-household social contacts to less than 3
2. I could reduce my social contacts to less than 5
3. I could reduce my social contacts to less than 10
4. I would not be willing to reduce my social contacts
5. I would not be able to reduce my social contacts

Base: all respondents

**Q13 [S]**

If there was a recommendation to get vaccinated during a future pandemic, how likely would you be to get vaccinated?

1. Very likely
2. Somewhat likely
3. Somewhat unlikely
4. Very unlikely
5. Not sure

Base: all respondents	QF1 [O]

Do you have any comments or clarifications about your answers? Please write them in the box below.

**[INSERT STANDARD CLOSE]**

37

### Appendix B: Regressions without top coding

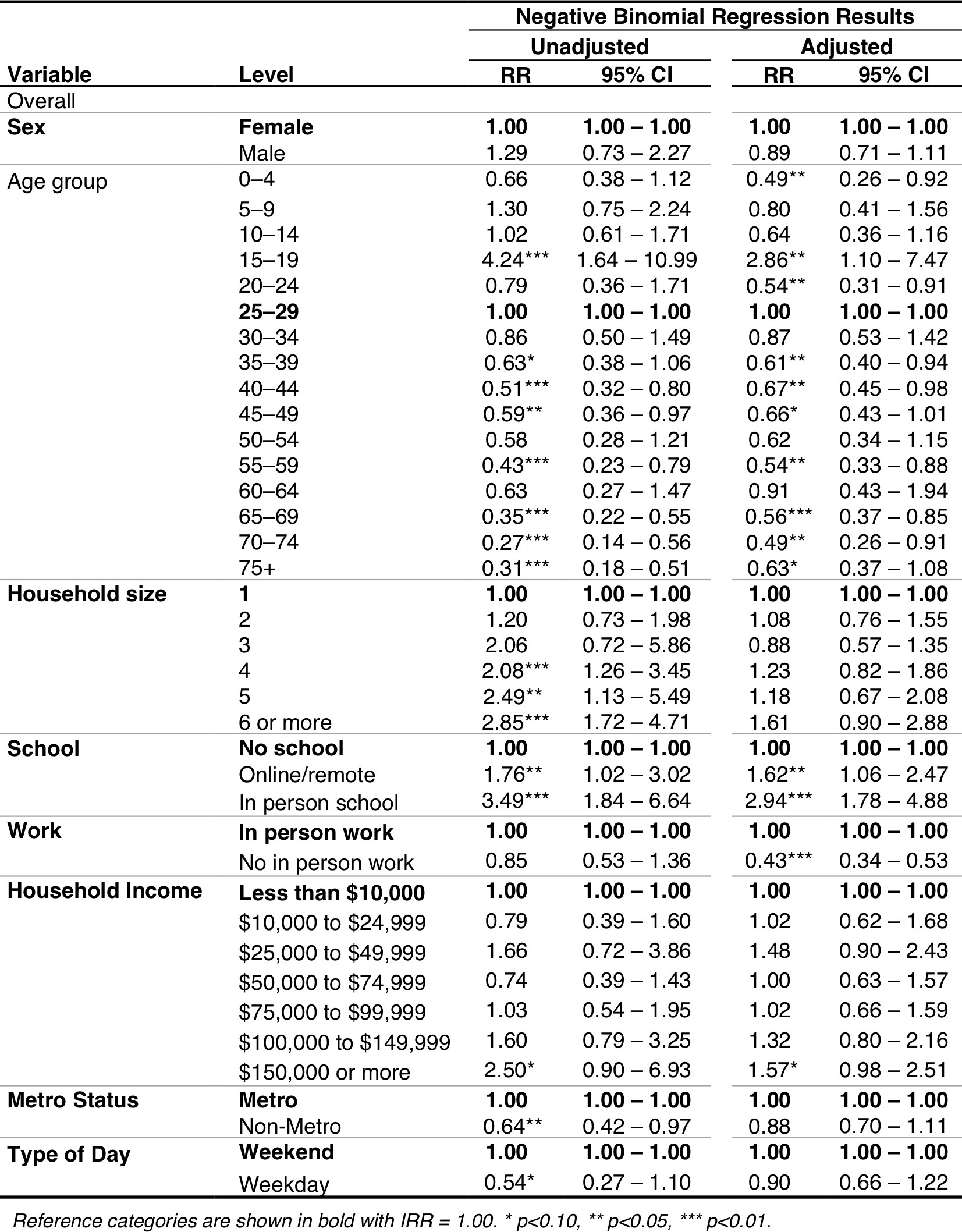
**Table B1**. Unadjusted and adjusted rate ratios (RR) for contact counts by sociodemographic factors, using participant’s reported total contacts, non-top coded.

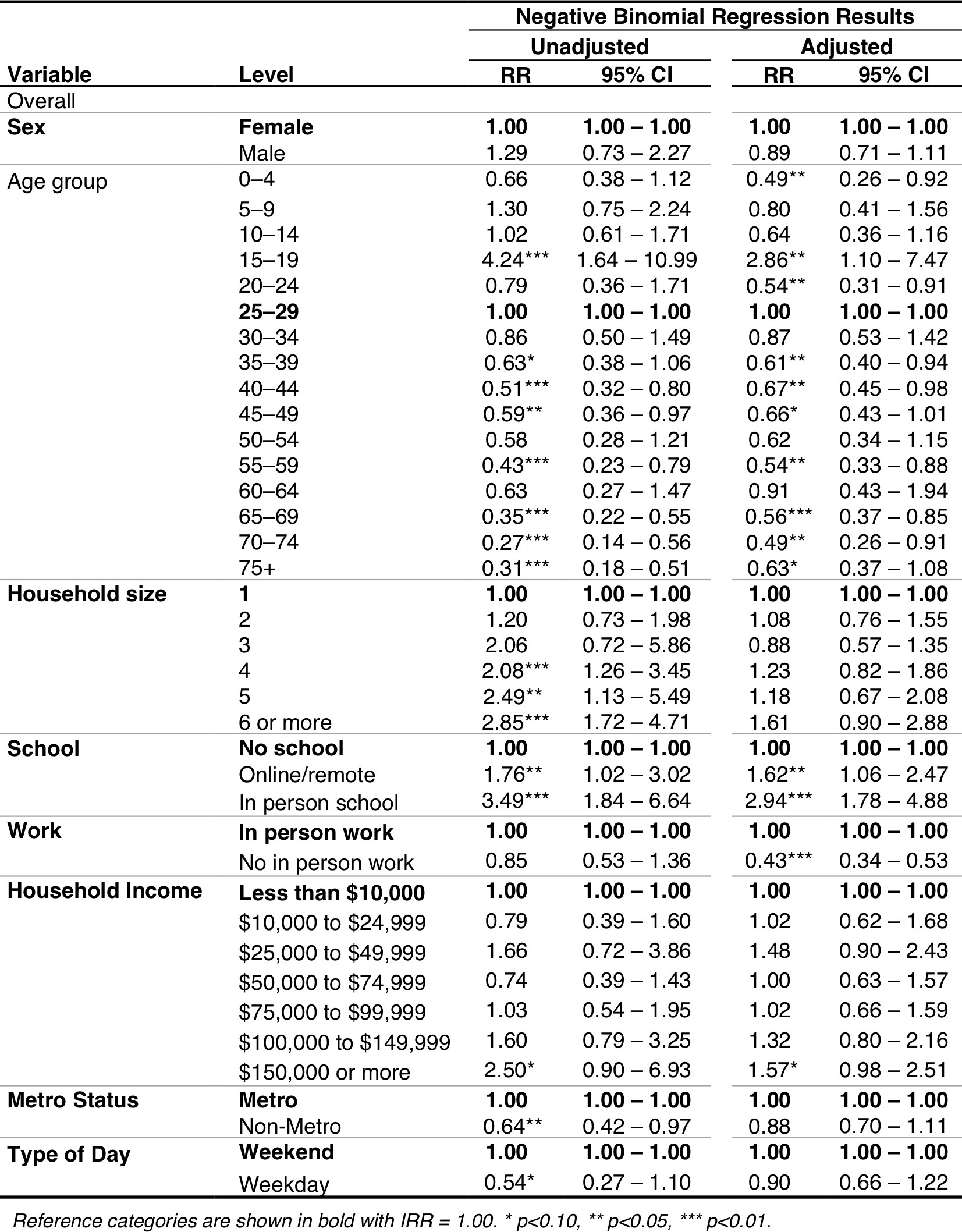

**Appendix C: Symmetric Contact Matrices**

Due to differences in reporting, the matrices that we generated in the main paper are not symmetric/reciprocal [16, 19]. This appendix contains reciprocal contact matrices (*M^k^* ) and their accompanying Q-indices. For reciprocal matrices at the population level, the total number of contacts reported between participants in age group *i* and contacts in age group *j* is equal in both directions. We use data from the 2023 American Community Survey to define *N_i_* and *N_j_*, the population sizes of age group *i* and *j*. The entries of the reciprocal

*ij*

matrices *M^k^*	were calculated as:

*ij*

$$M_{ij}^{k}=\frac{M_{ij}N_{i}+M_{ji}N_{j}}{\begin{aligned} 2N_{i} \\ \end{aligned}}$$

### **Figure C1.** Symmetric age-structured contact matrices by setting.

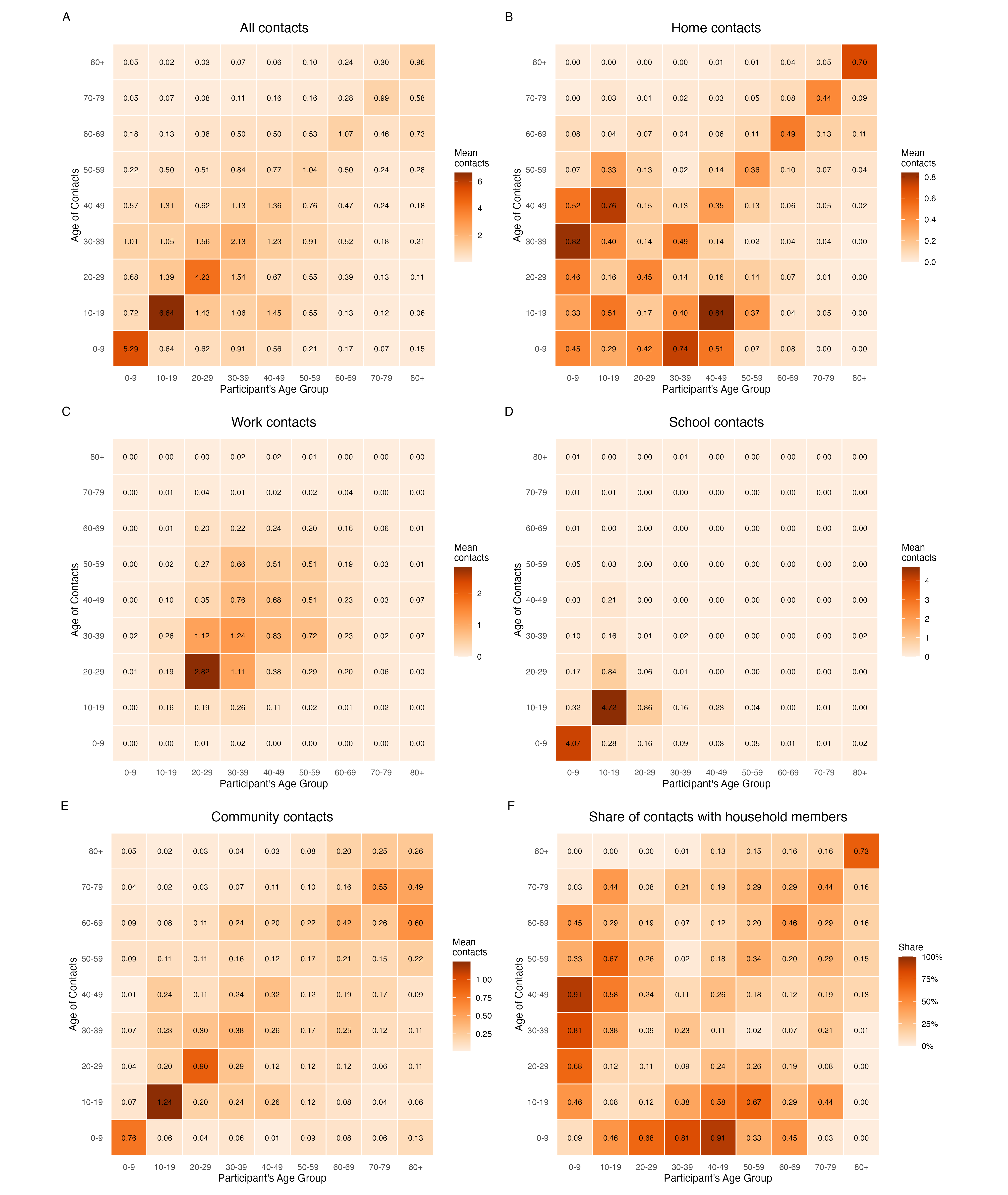

### Appendix D

**Figure D1.** Setting specific histograms. Maximum number of contacts is top-coded at 40.
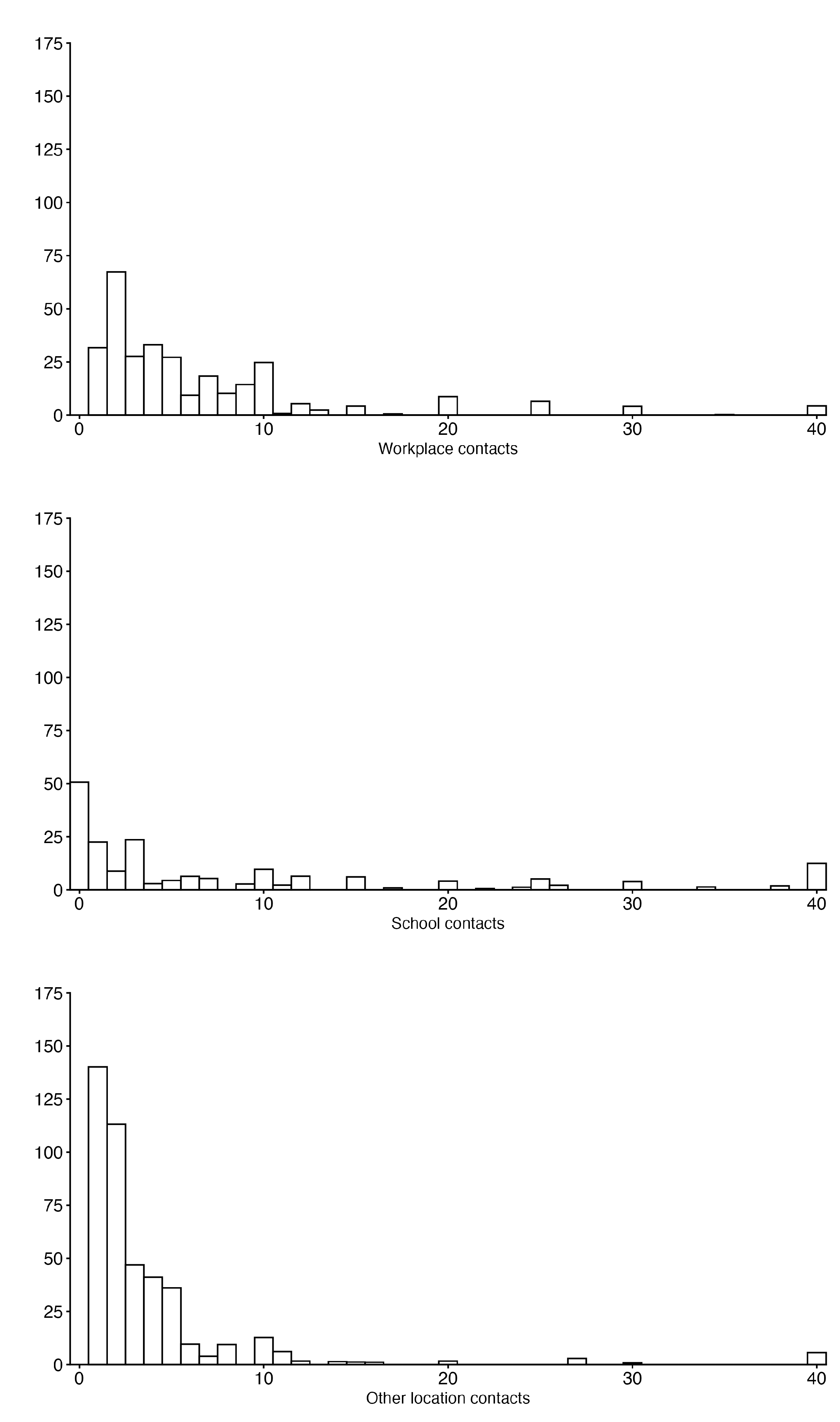

**Appendix E**

**Figure E1.** Histogram showing the distribution of the total number of daily contacts, without top-coding. Mean is 8.68 and median is 4.0.

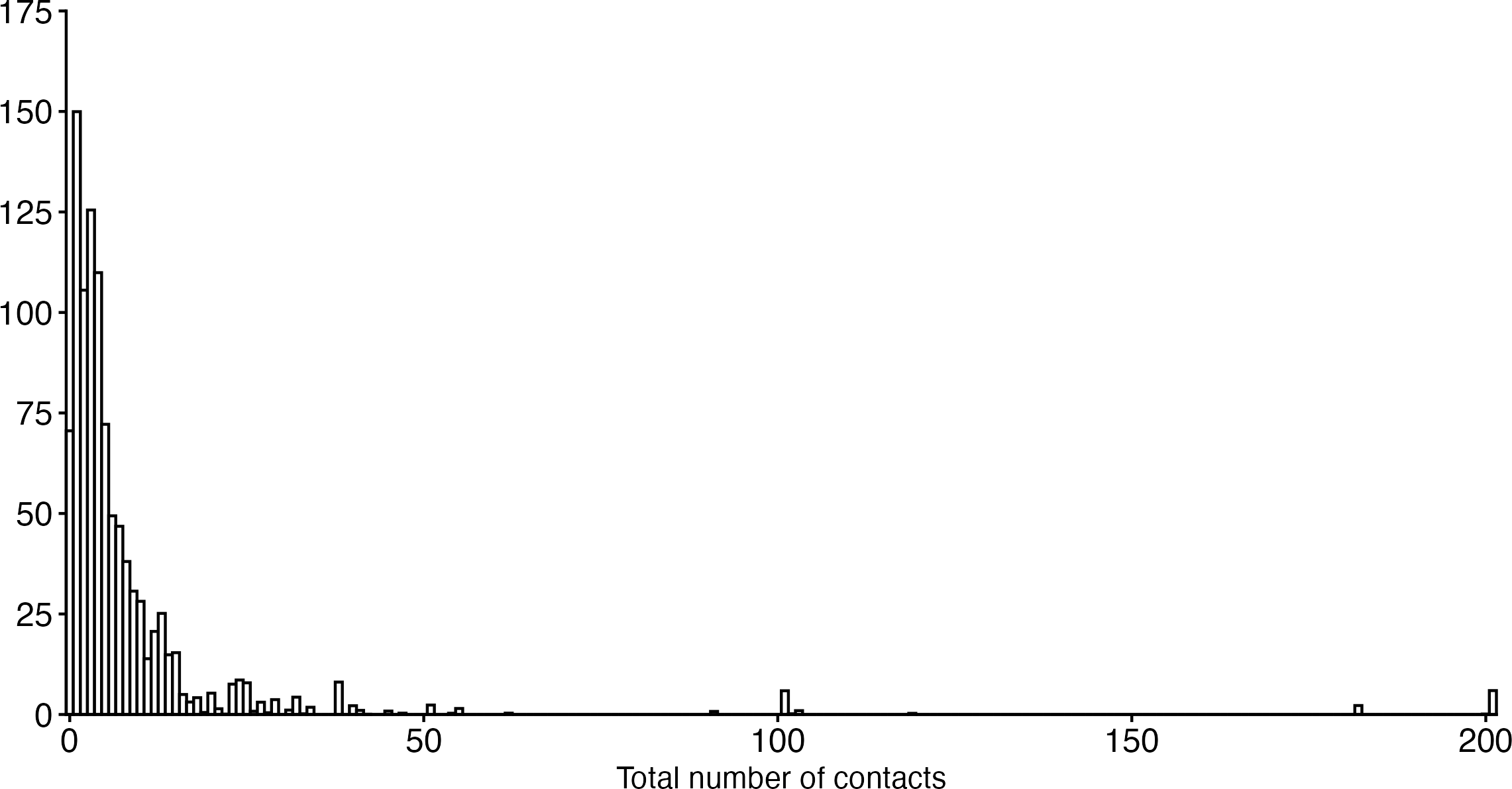
